# Neonatal fecal abundance of *Bifidobacterium longum* subspecies *infantis* is not associated with anthropometric outcomes up to 6 months of age in Bangladeshi infants

**DOI:** 10.1101/2025.07.07.25329844

**Authors:** Cole K. Heasley, Veselina Stefanova, Celine Funk, Aline C. Freitas, Grace Li, Lisa G. Pell, Diego G. Bassani, Karen M. O’Callaghan, Prakesh S. Shah, Jakaria Shawon, S.M. Abdul Gaffar, Rashidul Haque, Shafiqul Alam Sarker, Daniel E. Roth

**Affiliations:** Centre for Global Child Health, The Hospital for Sick Children, Toronto, Canada; Child Health Evaluative Sciences, The Hospital for Sick Children, Toronto, Canada; Dalla Lana School of Public Health, University of Toronto, Toronto, Canada; Department of Paediatrics, University of Toronto, Toronto, Canada; Department of Nutritional Sciences, King’s College London, London, United Kingdom; Department of Pediatrics, Mount Sinai Hospital, Toronto, Canada; Nutrition Research Division, International Centre for Diarrhoeal Disease Research, Bangladesh, Dhaka, Bangladesh; Infectious Diseases Division, International Centre for Diarrhoeal Disease Research, Bangladesh, Dhaka, Bangladesh

**Keywords:** *Bifidobacterium longum* subspecies *infantis*, Anthropometry, Growth, Child, Gastrointestinal microbiome, Microbiota, Infant health

## Abstract

*B. infantis* abundance in the infant gut may be associated with growth and health outcomes. However, these relationships have not been widely studied in settings where *B. infantis* is a dominant early-life commensal and growth faltering is prevalent. Here, we estimated associations between neonatal *B. infantis* abundance and anthropometric outcomes up to 6 months of age in generally healthy infants in Dhaka, Bangladesh; diarrhea and hospitalizations (at 1-2 and 6 months) were secondary morbidity outcomes. *B. infantis* stool absolute abundance was quantified by qPCR; for each infant, the primary exposure was mean abundance (0-28 days). Length-for-age, weight-for-age, and weight-for-length z-scores were derived at birth, 2, 3, and 6 months. Neonatal *B. infantis* abundance had a bimodal distribution, with 63% of infants having detectable *B. infantis* by 28 days of age. Anthropometric z-score distributions were shifted down, with means below zero at all ages. Neonatal *B. infantis* abundance was not associated with any anthropometric outcome at 2, 3, or 6 months of age (n=830), or with the risks of diarrhea or hospitalizations. The lack of association of neonatal *B. infantis* abundance with growth outcomes suggests that promoting early *B. infantis* colonization is unlikely to improve growth in populations with postnatal faltering.

## Introduction

*Bifidobacterium longum* ssp. *infantis* (*B. infantis*) is an early-life colonizer of the infant gut that is well adapted to metabolize human milk oligosaccharides (HMOs), a group of complex indigestible sugars present in human milk (Frese et al., 2017; Underwood et al., 2015). HMO metabolism by *B. infantis* produces lactic and acetic acids and other short-chain fatty acids that contribute to a lower pH in the gut lumen, inhibit proliferation of potential pathogens, and may influence immunoregulation and immune development (Frese et al., 2017; Fukuda et al., 2011; Henrick et al., 2021; Huda et al., 2019; Laursen et al., 2021). *B. infantis* colonization is hypothesized to improve infant health including linear and ponderal growth outcomes, possibly by reducing the risk of common morbidities such as diarrheal illness, and through effects of short-chain fatty acids on host metabolism including gluconeogenesis (Boets et al., 2017).

However, there is a lack of epidemiological evidence in support of an effect of early postnatal *B. infantis* colonization on infant growth. Few studies have been conducted in low- and middle-income countries (LMICs); in one study of infants in Bangladesh, there was a positive association between *B. infantis* and infant weight-for-age z-score (WAZ) and weight-for-length z-score (WLZ) in the first year after birth (Vatanen et al., 2022), but in a multivariable-adjusted analysis of cohorts in Bangladesh, Tanzania, and Pakistan, there were no associations between neonatal *B. infantis* abundance and length-for-age z-score (LAZ) or WAZ at 24 months of age (Colston et al., 2022). A key consideration in the interpretation of epidemiological studies is the wide global variation in *B. infantis* abundance – in general, it is rare or even absent among infants in high-income countries but frequently detected in LMICs (Lawley et al., 2019; Shao et al., 2025; Taft et al., 2022), settings where infant growth faltering is most common (UNICEF, 2023; Victora et al., 2021). Therefore, while *B. infantis* abundance might be found to explain between-individual variations in linear or ponderal growth within a study population, these associations may not explain why nearly all infants in LMICs grow more slowly than expected for their chronological age based on international growth standards (Roth et al., 2017).

In a cohort of generally healthy and mostly term-born infants in Dhaka, Bangladesh, we previously found *B. infantis* was highly abundant in stool by 2 months of age and persisted to at least 6 months of age; however, it was generally absent in the immediate postnatal period and there was substantial variation in abundance in the first month (neonatal period) (Freitas et al., 2025). To explore the potential role of postnatal *B. infantis* colonization in modulating growth in early infancy, we examined the associations between *B. infantis* abundance in the neonatal period and anthropometric outcomes up to 6 months of age in the same cohort. In further analyses, we estimated associations of *B. infantis* with stool pH and infant morbidity (diarrhea and hospitalizations), considering that such relationships might represent pathways by which *B. infantis* could influence infant growth.

## Methods

### Study design and participants

This secondary-use study was based on data from the Synbiotics for the Early Prevention of Severe Infections in Infants (SEPSiS) observational cohort study (ClinicalTrials.gov ID: NCT04012190). Approval for the original data collection was provided by The Hospital for Sick Children Research Ethics Board (REB# 1000063899) and the International Centre for Diarrhoeal Disease Research, Bangladesh (icddr,b) Research Ethics Committee (PR-19045). Approval for this secondary use study was obtained from The Hospital for Sick Children (REB#1000082019). Further details on the design and implementation of the SEPSiS observational cohort study have been described elsewhere (Freitas et al., 2025; Fung et al., 2025). Briefly, between November 2020 and February 2022, infants were enrolled at two government healthcare facilities in Dhaka, Bangladesh: Maternal and Child Health Training Institute and the Mohammadpur Fertility Services and Training Centre. Infants were eligible for enrolment if they were delivered at a study hospital, age zero (day of birth) to 4 days old, orally feeding at the time of screening, and their caregivers were planning to maintain residence in the study catchment area until the infant reached at least 60 days of age. Infants were ineligible for enrolment if they weighed <1500 g at birth, had a major congenital anomaly of the gastrointestinal tract, death or major surgery were likely within the first week after birth, were on mechanical ventilation or cardiac support, were receiving or prescribed parenteral antibiotics, were enrolled in an ongoing clinical trial involving the administration of probiotics and/or prebiotics, resided in the same household as another infant under 60 days of age (excluding twins), prenatal or postpartum/postnatal non-dietary probiotic supplements were used by the infant’s mother or provided directly to the infant, multiple gestation of the mother exceeded two, or the mother was diagnosed with HIV infection and/or history of receiving anti-retroviral drug(s) for a presumed HIV infection. For each model assessed, this study included all SEPSiS observational cohort participants who had relevant data.

### Stool sample collection

Study personnel collected infant stool samples using standardized procedures according to one of three schedules (A, B, or C). Infants in schedules A and B had up to 11 routine stool samples collected (days 0 or 1, 3 or 4, 6 or 7, 10, 14, 21, 28, 35, 60, 90, and 180), whereas infants in schedule C had up to 3 routine stool samples collected (on the day of enrollment and twice more on randomly assigned visits up to day 60). Infants on schedules A and B were combined in this analysis because they used the same stool sample collection schedules, had similar number of samples per infant, and had no other differences in the protocol that would influence the current analysis. In hospital or the infant’s home, stool samples were collected using a wrapped plastic sheet or plastic wrap-lined diaper, then manually homogenized and aliquoted into appropriate vials. For samples collected by study workers (all schedule A/B samples, 46% of schedule C samples), aliquots intended for qPCR were placed in the vapour phase of liquid nitrogen within 20 minutes of defecation, and aliquots for stool pH measurements were stored in a cold box at 2-8□C (never frozen). For schedule C samples that could not be collected directly by study workers (54% of samples), the infant’s caregiver was instructed to place the stool sample in a cold box (maintained at 2-8□C) within 20 minutes of defecation until it was transported to the field office by study staff. All stool aliquots were placed into the vapour phase of liquid nitrogen within 6 hours of defecation, before transport to icddr,b for storage at ≤-70□C until analysis.

### Sample processing and laboratory analyses

Stool sample selection and quantification of *B. infantis* absolute abundance have been described in detail elsewhere (Freitas et al., 2025). Briefly, total DNA was extracted from 100-150 mg of stool using QIAamp Fast DNA Stool Mini Kit (QIAGEN, Germany). Total DNA mass was quantified using a Nanodrop 2000 spectrophotometer (Thermo Scientific, USA). *B. infantis* was quantified using qPCR with primer-probes specific to *B. infantis* (Lawley et al., 2017). Cell counts were estimated using standard curves generated using pure cultures of *B. infantis* ATCC 15697, and absolute abundance was normalized to total DNA mass and expressed as log *B. infantis* cells per µg total DNA. For samples with undetectable *B. infantis* (*i.e.*, did not amplify or had cell counts below the limit of detection of the qPCR assay), a value of 2.5 log cells/µg DNA was imputed for the absolute abundance. This value corresponds to the median of half the limit of detection normalized to each undetected sample DNA mass.

Aliquots for measuring stool pH were brought to room temperature before mixing 1 g of stool with deionized water to achieve a total sample volume of 10 mL. Stool pH was measured twice per sample using portable pH testers (Hanna, USA); if the difference between the two pH measurements was greater than 0.5 pH units, a third measurement was obtained. The mean of the two pH values (or the mean of the two closest values if three were available) was considered the pH measurement of each stool sample used in further analyses described below.

### Clinical and anthropometric data collection

Routine study visits were conducted five times at the infant’s home or via phone call up to and including 14 days of age, then weekly until 60 days, then at 3 and 6 months. Study staff collected information about feeding practices (7-day recall), infant receipt of antibiotics (since last study visit), diarrheal episodes, and hospital admissions (Fung et al., 2025). Diarrheal episodes were based on a caregiver response of ‘yes’ when asked if the infant had diarrhea described as the “frequency of stool is too often, the quantity of stool is too much, and/or the consistency is too loose compared to the usual,” since the prior study visit or in the preceding 7 days, including the day of the encounter. Hospitalizations were verified and recorded by Study Medical Officers and were defined on the basis of admission to hospital for any reason other than injury or elective surgery.

Anthropometric measurement methods were adapted from standardized procedures from the INTERGROWTH-21^st^ study (Cheikh Ismail et al., 2013). Trained study staff measured infant length and weight on the day of enrolment (days 0-4) and during scheduled home visits at postnatal ages 2, 3, and 6 months. Length was measured using a wooden length board to the nearest 1 mm (Shorr board, Weight and Measure LLC, USA) and weight was measured with a Seca 334 or 354 digital scale to the nearest 1 g (Seca, USA). Anthropometric measurements were obtained twice and averaged; if the difference between two measurements was greater than 7 mm or 50 g for length and weight, respectively, a third measurement was taken. If a third measurement was taken, the two closest measurements were used to calculate the average.

For measurements on days 0-2 of age, baseline LAZ and WAZ were derived using INTERGROWTH-21^st^ Newborn Size growth curves (The International Fetal and Newborn Growth Consortium for the 21st Century, 2024a). For measurements obtained beyond 2 days of age, including at the 2, 3, and 6 months of age visits, the WHO Child Growth Standards were used to derive LAZ and WAZ for term infants (gestational age ≥259 days) (WHO, 2006), whereas for preterm infants (gestational age <259 days) older than 2 days postnatal age, LAZ and WAZ were generated using the INTERGROWTH-21^st^ Postnatal Growth for Preterm Infants growth curves (The International Fetal and Newborn Growth Consortium for the 21st Century, 2024b). Gestational age at birth was based on recalled last menstrual period (LMP) or prenatal ultrasound reports, following American College of Obstetricians and Gynecologists guidelines (“Committee Opinion No 700: Methods for Estimating the Due Date,” 2017). All WLZs were derived from WHO Child Growth Standards as the index is independent of age; baseline WLZ was based on concurrent weight and length measured at enrolment (days 0-4), but was not generated if the length was <45 cm. In instances of implausible values (infant length or weight decreasing over time or LAZ/WAZ ≥±6 or WLZ ≥±5), values and trajectories were examined independently by two reviewers to determine if any measurements should be removed. In total, 92 (4.8%) infants were flagged for investigation among whom 51 discrete length and 14 weight measurements were removed from analysis. For primary analyses, anthropometric z-scores were grouped according to the following age intervals: baseline (days 0-4), 2 months (days 59-75), 3 months (days 76-110), and 6 months (days 160-200).

### Statistical analysis

Baseline characteristics were described using medians (with 25^th^ and 75^th^ centiles) for continuous data and counts (proportions) for categorical data. Scatterplots with LOESS curves were generated for all *B. infantis*-anthropometric associations investigated to visualize trends in bivariate relationships.

*B. infantis* absolute abundance in the first 28 days after birth was summarized at the infant level using the mean of each infant’s qPCR abundance data from all analyzed samples collected from 0-28 days (neonatal period) and is referred to as ‘neonatal *B. infantis* absolute abundance’. Several summary measures of neonatal *B. infantis* absolute abundance were tested and the mean was selected for primary analyses (see Supplementary Methods, **Table S1 and Figures S1-3**). Associations between neonatal *B. infantis* absolute abundance and anthropometric indices at specified time points were estimated using linear regression models. All models with anthropometric outcomes included adjustment for the same anthropometric index measured at baseline. Sample sizes for the otherwise unadjusted models included only those infants with complete covariate data and who were included in the corresponding multivariable-adjusted analysis. Additional covariates in the adjusted regression models included: breastfeeding status assessed at or closest to age 28 days, infant sex, hospital enrolment site, mode of delivery, gestational age at birth, neonatal infant systemic antibiotics (ever/never), maternal BMI at enrolment, maternal education, maternal age, the number of infants within the household under 5 years of age, and household asset index quintile. Asset index was calculated for all infants enrolled in the SEPSiS observational cohort and a related clinical trial (Pell et al., 2025) using principal component analysis of a checklist of household assets and scored based on the first principal component, which was then converted to quintiles. The primary outcome was LAZ, and the secondary outcomes were WAZ and WLZ.

In sensitivity analyses, we used the following alternative approaches to define neonatal *B. infantis* abundance: 1) area under the curve (of absolute abundance by age, as a continuous variable); 2) *B. infantis* absolute abundance expressed as a categorical dichotomous variable defined as undetectable (all stool samples within the first 28 days of age with undetectable *B. infantis*) or detectable (at least one stool sample with detectable *B. infantis* within the first 28 days of age). We also conducted sensitivity analyses in which we modified the period during which *B. infantis* absolute abundance was summarized for each infant (mean of available data for 0-14 or 0-60 days of age), and a cross-sectional sensitivity analysis using the single absolute abundance measurement that was closest in time to the 2-month anthropometric measurement (and collected within the interval of 59-75 days). We repeated the primary analysis in a subset of the cohort restricted to infants assigned to specimen collection schedules A/B. Lastly, we estimated the association between baseline anthropometric indices and neonatal *B. infantis* abundance as the outcome, and we performed the primary analyses without adjustment for the baseline measures.

To estimate the cross-sectional association between *B. infantis* absolute abundance and stool pH (using corresponding values generated for the same stool sample), we used unadjusted linear mixed effects regression models including all stool samples collected in the neonatal period. Using the same modeling approach, we also compared the stool pH of samples with detectable versus undetectable *B. infantis*.

For each infant, ‘neonatal stool pH’ was defined as the median of their pH measurements among all stool samples collected from 0-28 days. Associations of neonatal stool pH with LAZ, WAZ, and WLZ at 2, 3, and 6 months were assessed using multivariable linear regression models with the same covariates as described above. In a sensitivity analysis, we conducted a cross-sectional analysis of the association of stool pH with anthropometric outcomes using the pH measurement of the single stool sample collected closest in time to each anthropometric measurement (and within the same age interval used for the corresponding anthropometric value).

To estimate associations of neonatal *B. infantis* abundance with diarrheal events from 29 to 60 days (2 months) of age and hospitalizations (excluding trauma/injury or elective surgeries) from 29 to 180 days (6 months) of age, infants were categorized as never/ever having had a diarrheal event or hospitalization during the defined period. Modified Poisson regression models (with robust standard errors) were used to estimate relative risks of diarrhea and hospitalization by neonatal *B. infantis* abundance. In adjusted models, covariates included breastfeeding status assessed on or closest to 28 days of age, mode of delivery, gestational age, neonatal antibiotic exposure, maternal education, and household asset index quintile.

We considered the use of path analysis to examine if any associations between *B. infantis* abundance and anthropometric indices could be explained by mediating roles of stool pH or clinical events, but these analyses were not pursued due to null associations of *B. infantis* abundance with anthropometric indices.

The Holm method was used to account for multiple comparisons and maintain an overall α of 0.05 for each set of analyses (one set refers to all analyses presented within each table). All analyses were conducted using R statistical software (Version 4.4.1) (R Core Team, 2024). All statistical code used in analyses for this study was independently reviewed by a second data analyst.

## Results

Of 1939 infants enrolled in the SEPSiS observational cohort, 966 infants contributed 3903 stool samples in the neonatal period, of which 2982 (76%) underwent *B. infantis* qPCR testing. From 0-28 days of age, infants assigned to schedules A/B and C contributed a median (25^th^, 75^th^ centiles) of 6.0 (2.0, 7.0) and 1.0 (1.0, 3.0) stool samples for qPCR analysis, respectively. There were 830 infants (43% of 1939 enrolled) with at least one *B. infantis* qPCR measurement (0-28 days), at least one anthropometric outcome measure (2, 3, and 6 months) and a corresponding baseline anthropometric measure, and complete covariates, who were therefore included in at least one of the primary analyses of associations of *B. infantis* abundance and anthropometric outcomes (**Figure 1**). Eligible infants were predominantly born full-term (93%), via C-section (52%), and 68% were exclusively breastfed up to 28 days of age (**Table 1, Table S2** stratified by infant sex). There were 1072 infants (55%) with at least one stool pH measurement in the neonatal period, at least one anthropometric outcome measurement in one of the age intervals assessed and complete covariates, who were therefore included in at least one of the primary analyses of associations of stool pH and anthropometric outcomes (**Figure 1**). There were 938 and 935 infants (48%) with a neonatal *B. infantis* absolute abundance measurement who also had morbidity-related data up to 2 months or hospitalization data up to 6 months of age, respectively.

**Figure 1.**
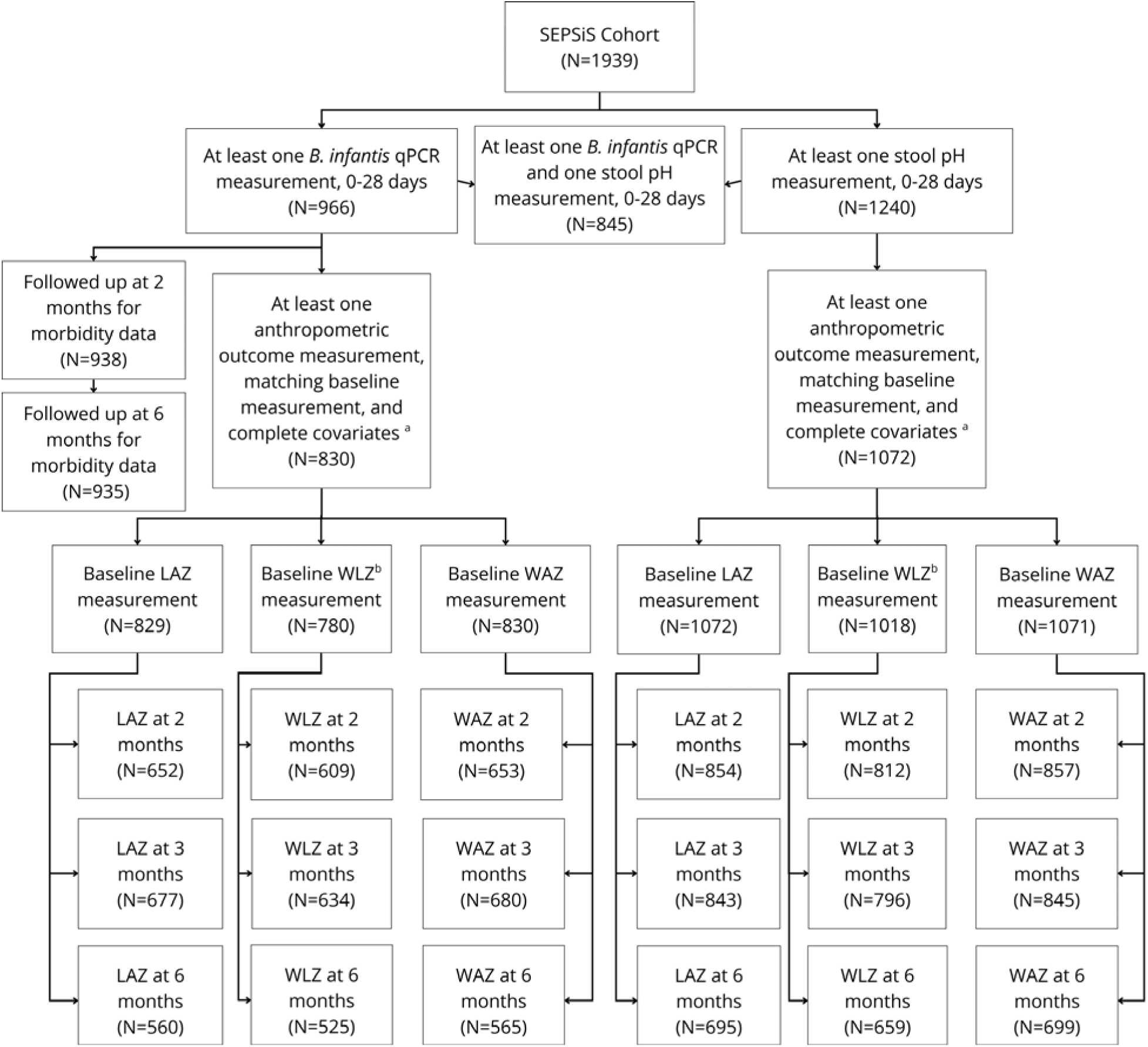
Study flow diagram. Numbers of infants from the SEPSiS observational study cohort who contributed to the primary and secondary analyses in this study. ^a^ Complete covariates include: enrolment site, infant sex, breastfeeding status at 28 days of age (if available) or at age most proximal to 28 days, mode of delivery, gestational age at birth, infant antibiotic exposure by 28 days of age, maternal BMI, maternal education, maternal age, number of infants within the household under 5 years of age, and household asset index quintile. ^b^ Sample sizes for WLZ were lower than for the corresponding LAZ and WAZ datasets because WLZ was derived using the WHO child growth standards, whereby WLZ was not generated for some baseline measurements if length and weight were not measured at the same visit, or when length was <45cm; however the INTERGROWTH-21^st^ standards enabled baseline LAZ to be generated for newborn lengths below 45 cm.

**Table 1.** Baseline characteristics of infants included in primary analyses of the associations of neonatal *B. infantis* absolute abundance and anthropometric outcomes.

| Characteristic | Median (25<sup>th</sup>, 75<sup>th</sup> centiles) or count (%)<br>(n=830) |
| --- | --- |
| Enrolment site |  |
| Mohammadpur Fertility Services and Training Centre | 477 (57) |
| Maternal and Child Health Training Institute | 353 (43) |
| Sex |  |
| Male | 409 (49) |
| Female | 421 (51) |
| Baseline LAZ<sup>a, b</sup> | -0.83 (-1.5, -0.15) |
| Baseline WAZ<sup>a, b</sup> | -1.0 (-1.7, -0.4) |
| Baseline WLZ<sup>a, b</sup> | -0.61 (-1.4, 0.16) |
| Birth weight<sup>b</sup>, kg | 2.87 (2.60, 3.11) |
| Mode of delivery |  |
| Vaginal | 400 (48) |
| C-section | 430 (52) |
| Stool collection schedule<sup>c</sup> |  |
| A/B | 320 (39) |
| C | 510 (61) |
| Breastfeeding status by 28 days of age |  |
| Exclusively breastfed | 561 (68) |
| Predominantly breastfed | 157 (19) |
| Partially breastfed or not breastfed | 112 (13) |
| Gestational age at birth |  |
| Term ($\geq 259$ days) | 771 (93) |
| Preterm ($< 259$ days) | 59 (7.1) |
| Antibiotic use by 28 days of age |  |
| Yes | 94 (11) |
| No | 736 (89) |
| Maternal BMI, kg/m<sup>2</sup> | 26 (23, 29) |
| Maternal education |  |
| Little to no schooling<sup>d</sup> | 225 (27) |
| Secondary incomplete | 305 (37) |
| Secondary complete or higher | 300 (36) |
| Maternal age, years | 24 (20, 28) |
| Infants within the household under 5 years of age | 0 (0, 1) |
| Household asset index quintile<sup>e</sup> |  |
| 1 (lowest wealth) | 153 (18) |
| 2 | 168 (20) |
| 3 | 166 (20) |
| 4 | 179 (22) |
| 5 (highest wealth) | 164 (20) |
| Stool *B. infantis* AA measurements per infant, 0-28 days of age |  |
| Infants with 1 sample | 346 (42) |
| Infants with 2-4 samples | 189 (23) |
| Infants with 5-7 samples | 295 (36) |

Abbreviations: BMI, body mass index; LAZ, length-for-age z-score; WAZ, weight-for-age z-score; WLZ, weight-for-length z-score; qPCR, quantitative polymerase chain reaction; AA, absolute abundance.
<sup>a</sup> Mean and SD of anthropometric z-score distributions are shown in Table S3.
<sup>b</sup> $n < 830$ : baseline LAZ, $n=829$ ; baseline WLZ, $n=780$ . Baseline WAZ ( $n=830$ ) and WLZ were calculated from weights measured at enrolment by study staff, whereas birthweights ( $n=830$ ) were extracted from the hospital records.
<sup>c</sup> Infants in schedule A/B had up to 7 routine stool samples collected in the first month of age (days 0 or 1, 3 or 4, 6 or 7, 10, 14, 21, and 28). Infants in schedule C had up to 3 routine stool samples collected (day of enrolment and twice more on randomly assigned visits up to 60 days of age).
<sup>d</sup> Little to no schooling was defined as: no schooling at all; having completed *qawmi madrasa* but no other schooling; or, up to 5 years of schooling or *alia madrasa*.
<sup>e</sup> Quintiles of a score derived using the first component of principal component analysis from a checklist of household assets from all SEPSiS infants (observational cohort and associated trial). Higher quintile corresponds to higher asset ownership.

The distribution of neonatal *B. infantis* absolute abundances was bimodal (N infants = 830), with a median (25^th^,75^th^ percentiles) of 3.7 log cells/µg DNA (2.5, 8.8) (**Figure 2**). When the age range was reduced to 0-14 days, this median reduced to 2.9 (2.5, 6.9) log cells/µg DNA and when expanded to 0-60 days, it increased to 5.4 (2.5, 8.2) log cells/µg DNA. Among the 830 infants in the primary analyses, 524 (63%) had *B. infantis* detected in at least one stool sample. The median (25^th^, 75^th^) neonatal abundance among infants with detectable *B. infantis* was 6.4 log cells/µg DNA (2.5, 8.8); conversely, among infants with undetectable *B. infantis* in all stool samples from 0-28 days of age (37%), neonatal *B. infantis* abundances were all uniformly 2.5 log cells/µg DNA (corresponding to the imputed lower limit of quantification). The mean (SD) neonatal stool pH (n=1072 infants) was 5.7 (0.6).

**Figure 2.**
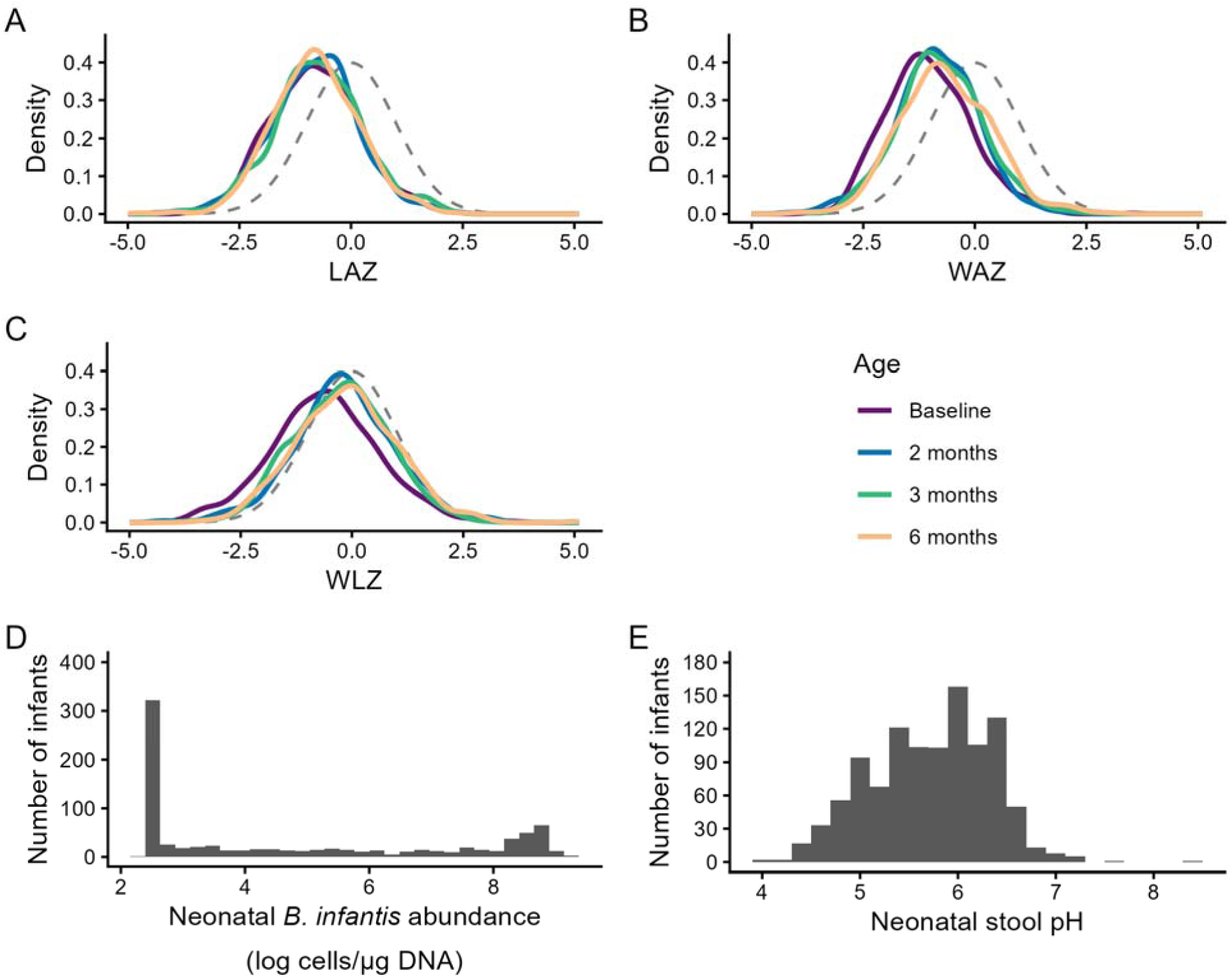
Distributions of anthropometric indices, neonatal *B. infantis* absolute abundance, and neonatal stool pH. Kernel density plots for infants with at least one *B. infantis* qPCR measurement and complete covariates: (**A**) LAZ (Baseline, n=829; 2 months, n=652; 3 months, n=677; 6 months, n=560), (**B**) WAZ (Baseline, n=830; 2 months, n=653; 3 months, n=680; 6 months, n=565), and (**C**) WLZ (Baseline, n=780; 2 months, n=609; 3 months, n=634; 6 months, n=525). Standard normal distributions (mean=0, SD=1) are shown as dashed grey lines in each panel. Histograms of (**D**) neonatal *B. infantis* absolute abundance (n=830) and (**E**) neonatal stool pH (n=1072). Abbreviations: LAZ, length-for-age z-score; WAZ, weight-for-age z-score; WLZ, weight-for-length z-score.

Anthropometric z-scores were approximately normally distributed with means below zero at all timepoints (**Figure 2** and **Table S3**). Infants included in this sub-study were generally representative of the overall SEPSiS observational cohort with respect to growth patterns (**Table S3**).

There were no associations between neonatal *B. infantis* absolute abundance and LAZ, WAZ, or WLZ, at 2, 3, or 6 months of age (**Table 2**; **Figure S4**). Neonatal *B. infantis* absolute abundance was not associated with baseline LAZ, WAZ, or WLZ (**Table S4**). Inferences remained unchanged when participants were stratified by sex (**Table S5)**, and in sensitivity analyses (**Tables S6-11**) with these exceptions: in the analysis restricted to infants enrolled in schedules A/B, there was a significant positive association between neonatal *B. infantis* abundance and LAZ at 3 months of age and a significant negative association between *B. infantis* and WLZ at 2 months of age, but both associations were attenuated and no longer significant at 6 months of age (**Table S12**). To investigate if the higher number of samples in schedule A/B may have influenced the associations, the analysis was rerun using two random samples per infant to resemble total sample numbers in schedule C, with selection simulated 1000 times. Based on this further analysis, we did not find evidence that the greater number of stool samples per infant in schedules A/B during the 0-28 day period explained the discrepancy in findings between the sub-analysis of schedule A/B and the primary analysis (**Table S13**).

**Table 2.**
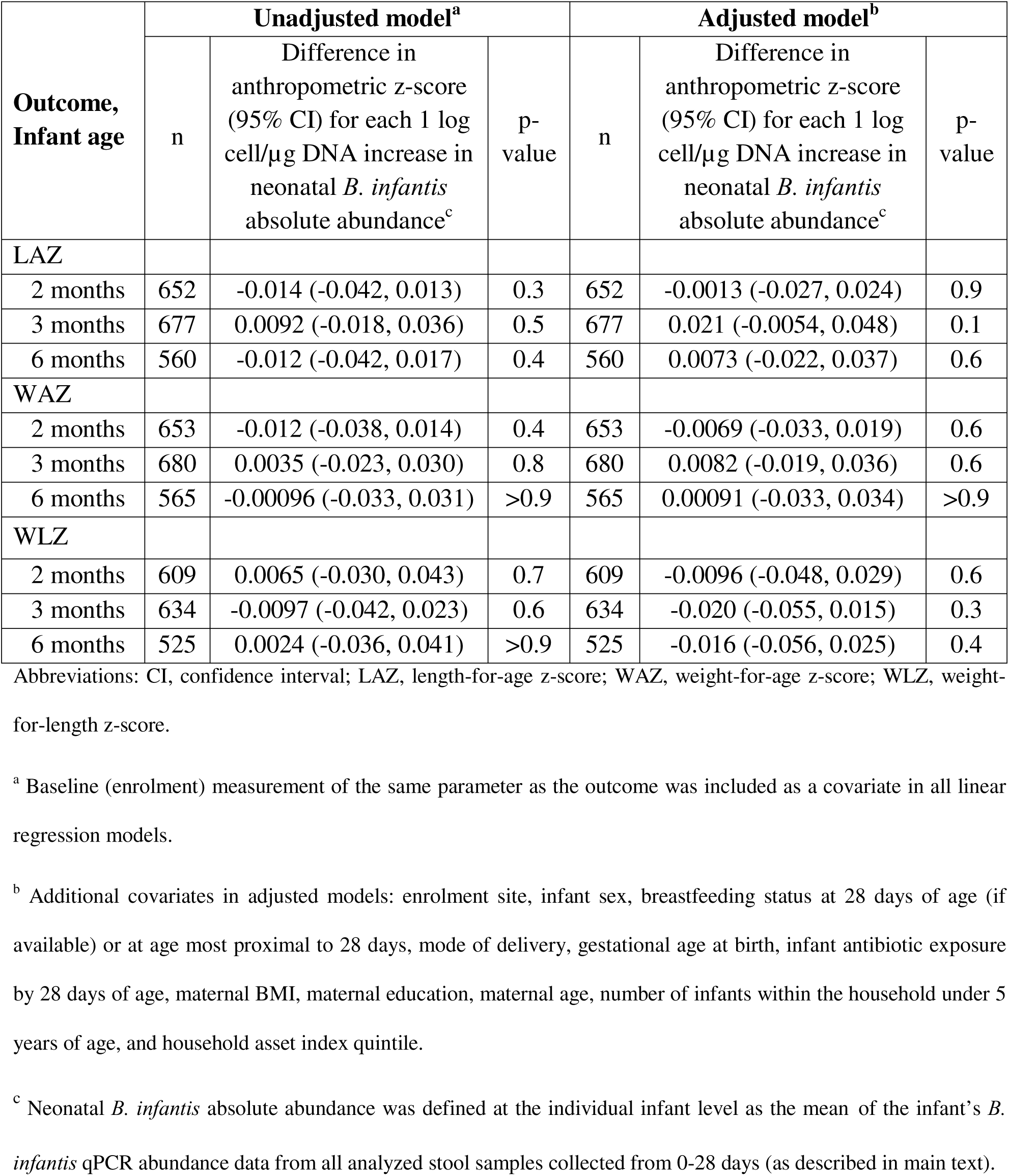
Associations of neonatal *B. infantis* absolute abundance (0-28 days of age) and infant anthropometric indices at 2, 3, and 6 months of age.

| Outcome,<br>Infant age | Unadjusted model<sup>a</sup> |  |  | Adjusted model<sup>b</sup> |  |  |
| --- | --- | --- | --- | --- | --- | --- |
| | n | Difference in anthropometric z-score (95% CI) for each 1 log cell/μg DNA increase in neonatal *B. infantis* absolute abundance<sup>c</sup> | p-value | n | Difference in anthropometric z-score (95% CI) for each 1 log cell/μg DNA increase in neonatal *B. infantis* absolute abundance<sup>c</sup> | p-value |
| LAZ |  |  |  |  |  |  |
| 2 months | 652 | -0.014 (-0.042, 0.013) | 0.3 | 652 | -0.0013 (-0.027, 0.024) | 0.9 |
| 3 months | 677 | 0.0092 (-0.018, 0.036) | 0.5 | 677 | 0.021 (-0.0054, 0.048) | 0.1 |
| 6 months | 560 | -0.012 (-0.042, 0.017) | 0.4 | 560 | 0.0073 (-0.022, 0.037) | 0.6 |
| WAZ |  |  |  |  |  |  |
| 2 months | 653 | -0.012 (-0.038, 0.014) | 0.4 | 653 | -0.0069 (-0.033, 0.019) | 0.6 |
| 3 months | 680 | 0.0035 (-0.023, 0.030) | 0.8 | 680 | 0.0082 (-0.019, 0.036) | 0.6 |
| 6 months | 565 | -0.00096 (-0.033, 0.031) | >0.9 | 565 | 0.00091 (-0.033, 0.034) | >0.9 |
| WLZ |  |  |  |  |  |  |
| 2 months | 609 | 0.0065 (-0.030, 0.043) | 0.7 | 609 | -0.0096 (-0.048, 0.029) | 0.6 |
| 3 months | 634 | -0.0097 (-0.042, 0.023) | 0.6 | 634 | -0.020 (-0.055, 0.015) | 0.3 |
| 6 months | 525 | 0.0024 (-0.036, 0.041) | >0.9 | 525 | -0.016 (-0.056, 0.025) | 0.4 |

Abbreviations: CI, confidence interval; LAZ, length-for-age z-score; WAZ, weight-for-age z-score; WLZ, weight-for-length z-score.
<sup>a</sup> Baseline (enrolment) measurement of the same parameter as the outcome was included as a covariate in all linear regression models.
<sup>b</sup> Additional covariates in adjusted models: enrolment site, infant sex, breastfeeding status at 28 days of age (if available) or at age most proximal to 28 days, mode of delivery, gestational age at birth, infant antibiotic exposure by 28 days of age, maternal BMI, maternal education, maternal age, number of infants within the household under 5 years of age, and household asset index quintile.
<sup>c</sup> Neonatal *B. infantis* absolute abundance was defined at the individual infant level as the mean of the infant's *B. infantis* qPCR abundance data from all analyzed stool samples collected from 0-28 days (as described in main text).

In individual neonatal stool samples (n=2118 samples), there was a 0.093 (95% CI: 0.084, 0.10; p <0.001) decrease in stool pH per one unit increase in log *B. infantis* cells/μg of DNA. There was evidence that the relationship between *B. infantis* absolute abundance with stool pH may be non-linear (**Figure S5**). When *B. infantis* abundance was dichotomized, the mean (SD) pH of stool samples with detectable and undetectable *B. infantis* was 5.5 (0.68) and 6.0 (0.54), respectively (p<0.001).

Neonatal stool pH up to 28 days after birth was significantly associated with WAZ at 3 months and WLZ at 2 and 3 months of age in unadjusted models (**Table 3** and **Figure S6**), but none of the associations were statistically significant after covariate adjustment and accounting for multiple testing (**Table 3**). There was no association between neonatal stool pH and LAZ at any timepoint (**Table 3**). Stool pH was not associated with any anthropometric indices in cross-sectional analyses (**Table S14**).

**Table 3.**
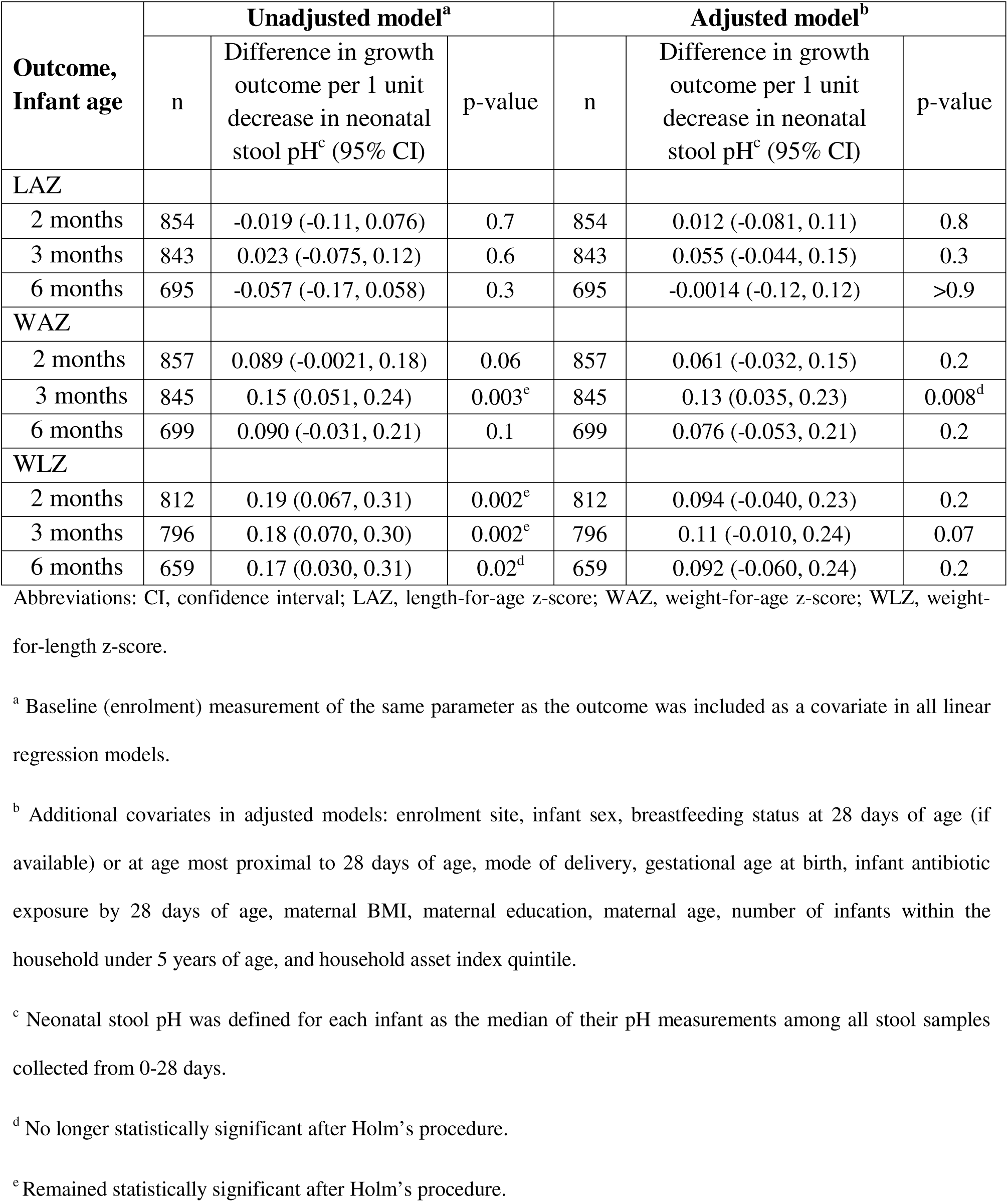
Associations of neonatal stool pH (0-28 days of age) and infant anthropometric indices at 2, 3, and 6 months of age.

Of the infants who had available *B. infantis* absolute abundance data and had not exited the study by the end of 2 or 6 months, 36 (3.8%) infants had at least one diarrheal event between 29 and 60 days of age (n=938), and 78 (8.3%) had at least one hospitalization event between 29 and 180 days of age (n=935). Neonatal *B. infantis* absolute abundance was not associated with the risk of an infant experiencing a diarrheal event (adjusted RR = 0.96 (95%CI: 0.82, 1.1) per 1 log *B. infantis* cells/μg of DNA increase; p = 0.5) or hospitalization (adjusted RR = 1.0 (95%CI: 0.90, 1.1) per 1 log *B. infantis* cells/μg of DNA increase; p>0.9).

## Discussion

In a cohort of generally healthy infants in Dhaka, Bangladesh, *B. infantis* stool absolute abundance in the neonatal period was not associated with anthropometric indices up to 6 months of age. Stool pH was inversely associated with stool *B. infantis* abundance, consistent with previous observations (Frese et al., 2017), but our results did not corroborate prior findings of an inverse association between stool pH and LAZ in Bangladeshi infants (Hossain et al., 2019). Although there was weak evidence of an inverse association of stool pH with infant WAZ in early infancy, the magnitude of the association was small given the narrow range of observed pH values, the association was not significant after accounting for multiple testing, and it was not sustained to 6 months of age. Additionally, this relationship could not be explained by a pH-lowering effect of *B. infantis* given the absence of an association of WAZ with stool *B. infantis* abundance; therefore, if the relationship between stool pH and weight represents a causal pathway, it may be attributable to other components of the microbiome, or it may be confounded by unmeasured aspects of the diet or medications that influence both stool pH and weight gain (Yamamura et al., 2023).

The null associations between *B. infantis* abundance and anthropometric indices in the first 6 months postnatal age were broadly consistent with a prior study that did not find relationships between neonatal *B. infantis* abundance and LAZ (n=292) or WAZ (n=472) at 24 months of age in Bangladesh, Pakistan, and Tanzania (Colston et al., 2022). In those Bangladeshi and Tanzanian cohorts, there was a positive association of enrolment (*i.e.*, birth) LAZ with *B. infantis* abundance (Colston et al., 2022), which raises the possibility of reverse causality. However, we did not observe a similar association in the SEPSiS observational cohort, and our inferences were the same in models with and without adjustment for baseline LAZ.

Recent systematic reviews of the effects of probiotics in infants have included few studies of *B. infantis*, or when included, were generally administered in combination with other probiotic species (Catania et al., 2021; Guo et al., 2023; Panchal et al., 2023), or focused on *B. infantis* but did not address growth outcomes (Batta et al., 2023). Two trials of the *B. infantis* probiotic EVC001 conducted in the United States did not find effects on achieved weight/weight gain or length among term exclusively breastfed infants (Smilowitz et al., 2017) or preterm infants (Bajorek et al., 2022). Similarly, two trials in Spain showed no effects of *B. infantis* probiotics on infant weight or length (Escribano et al., 2018; Manzano et al., 2017). However, in a single-blind randomized controlled trial involving 2- to 6-month old infants with severe acute malnutrition in Dhaka, Bangladesh (n= 67), infants treated with *B. infantis* EVC001 daily for 28 days had a significantly higher WAZ and mid-upper arm circumference compared to the placebo group at 28 days post-intervention, despite no effect on LAZ (Barratt et al., 2022; Nuzhat et al., 2023). Some studies of probiotics with multiple species including *B. infantis* reported positive effects on weight gain in very preterm infants in Australia and New Zealand (Jacobs et al., 2013) or very low birth weight infants in the United States (Al-Hosni et al., 2012) and Germany (Härtel et al., 2017), but it is uncertain whether the effects were specifically attributable to *B. infantis*. In a sensitivity analysis in the present study, which was restricted to a subset of infants who had stool sampled more frequently (schedule A/B; 40-42% of infants in primary analyses), *B. infantis* was positively associated with LAZ and negatively associated with WLZ at 3 months of age but these associations were substantially attenuated by 6 months of age and were not explained by a greater precision of *B. infantis* abundance measurement due to higher sample numbers. Therefore, we did not consider these findings to have altered the overall conclusion that neonatal *B. infantis* abundance is unrelated to growth in the first 6 months postnatal age.

A meta-analysis of trials investigating the effect of probiotics containing *B. infantis* on preterm and/or very low birth weight infants found that administering probiotics reduced all-cause mortality (14 trials with 4292 infants), necrotizing enterocolitis (15 trials with 3626 infants), and late onset sepsis (13 trials with 4123 infants) (Batta et al., 2023). However, only 5 of the 16 studies included 100 or more infants in each trial arm and all studies used a combination of probiotics that included *B. infantis*, rather than *B. infantis* alone. In the present study, we did not find associations between *B. infantis* neonatal abundance and either diarrheal events or hospitalizations; however, we acknowledge that these analyses may have been underpowered due to a low number of events. We did not assess associations with cause-specific hospitalizations due to the low number of events related to discrete causes. However, we acknowledge that the use of all-cause hospitalization as the outcome measure may have biased the association towards the null if only certain illnesses leading to hospitalization were truly associated with *B. infantis* abundance.

Examining individual-level risk factors, such as *B. infantis* abundance, may identify sources of inter-individual variations in growth within a population; however, inter-individual variations may not provide insights into a population-level phenomenon (Rose, 1985; Roth et al., 2017). Growth faltering in LMICs is not restricted to a subset of infants with stunting within a population, but rather the whole population distributions of anthropometric indices are shifted downwards (Roth et al., 2017; Victora et al., 2021), as observed in this study cohort. Given the high prevalence of *B. infantis* colonization in populations with endemic postnatal growth faltering (Lawley et al., 2019; Shao et al., 2025; Taft et al., 2022), and the absence of associations of *B. infantis* with growth at the infant level as observed here, there is unlikely to be an important role for early postnatal *B. infantis* colonization in mitigating growth-faltering in such settings.

Several limitations of the present study should be considered. Stool *B. infantis* abundance was used as a proxy of intestinal *B. infantis* abundance but stool abundance of specific microbes may not reflect abundance within the intestinal lumen (Zmora et al., 2018); however, methods for obtaining luminal samples from infants’ intestinal tracts rely on procedures that would be invasive, infeasible and unethical in a large cohort study in this age group. We focused our analysis on *B. infantis* colonization in the neonatal period because most infants were eventually colonized with *B. infantis* by 1-2 months of age (Freitas et al., 2025); therefore, we were unable to examine the effects of longer-term colonization. Additionally, we only explored the associations between infant growth and the abundance of naturally occurring *B. infantis* without consideration of strain-level distinctions. Beneficial effects from commensals or probiotics may be strain-specific; for example, HMO gene clusters are highly conserved across *B. infantis* strains, but the presence of specific genes within clusters differ among *B. infantis* strains (Duar et al., 2020; Locascio et al., 2010; Shao et al., 2025). The study population had a high C-section rate, which influences early gut microbial succession patterns (Freitas et al., 2025); however, this rate of C-section is comparable to other studies in this setting (National Institute of Population Research and Training (NIPORT) and ICF, 2022; Nuzhat et al., 2023). Furthermore, as in any observational study, the estimates should not be interpreted as causal effects as they may be biased by residual confounding due to a wide range of unmeasured factors related to the infant’s health status, components of the infant diet and microbiome composition.

## Conclusion

Stool absolute abundance of *B. infantis* in the first month after birth was not associated with linear or ponderal growth up to 6 months of age in generally healthy infants in Dhaka, Bangladesh. Based on these findings, promotion of early *B. infantis* colonization through interventions such as *B. infantis* probiotic administration in the neonatal period may not be expected to improve growth outcomes in generally healthy infants born in settings with high prevalence of natural *B. infantis* colonization and where postnatal growth faltering is widespread.

## Data Availability

All data produced are available online at

https://doi.org/10.5683/SP3/PWPMGK

## Acknowledgements

We would like to thank the families who participated in the SEPSiS cohort study, as well as the other SEPSiS project co-investigators and study personnel who were involved in data collection and laboratory analysis.

## Author Contributions

Conceptualization, C.K.H., V.S., L.G.P., D.G.B., K.M.OC., P.S.S., and D.E.R.; Methodology, C.K.H. and D.E.R.; Formal Analysis, C.K.H.; Data Curation, C.K.H., C.F., A.C.F., and G.L.; Validation, C.F., A.C.F., and G.L.; Writing—Original Draft, C.K.H., and V.S.; Writing—Review and Editing, C.K.H., V.S., C.F., A.C.F., G.L., L.G.P., D.G.B., K.M.OC., P.S.S., J.S., S.M.A.G., R.H., S.A.S., and D.E.R.; Supervision, D.E.R.; Funding Acquisition, D.E.R.

## Research Transparency and Reproducibility

All data and code used for the analyses are available on Borealis: https://doi.org/10.5683/SP3/PWPMGK

