## Supplement for "Neonatal fecal abundance of *Bifidobacterium longum* subspecies *infantis* is not associated with anthropometric outcomes up to 6 months of age in Bangladeshi infants"

*Current affiliation: Institute of Epidemiology, Disease Control, and Research, Dhaka, Bangladesh

### Supplemental Methods: Comparison of summary measures of neonatal *B. infantis* absolute abundance.

We assessed multiple methods of summarizing neonatal *B. infantis* absolute abundance at the infant level including the area under the curve (AUC), median, and mean. Other methods to summarize *B. infantis* absolute abundance, such as trajectory modelling, were considered but deemed unsuitable for our analyses because nearly half (44%) of the infants only had one sample measured in the neonatal period.

The calculation of an infant's *B. infantis* abundance AUC, using the AUC() function in R's DescTools package (version 0.99.58), is demonstrated here using two hypothetical examples. Both infants A and B had three *B. infantis* absolute abundance measures: 2.5 log cells/µg DNA, 4.5 log cells/µg DNA, and 8.0 log cells/µg DNA. Infant A had stool collected on 4, 21, and 25 days of age whereas infant B had stool collected on 4, 8, and 25 days of age (**Figure S1**). AUC was divided by 100 in order to rescale the variable for clarity.

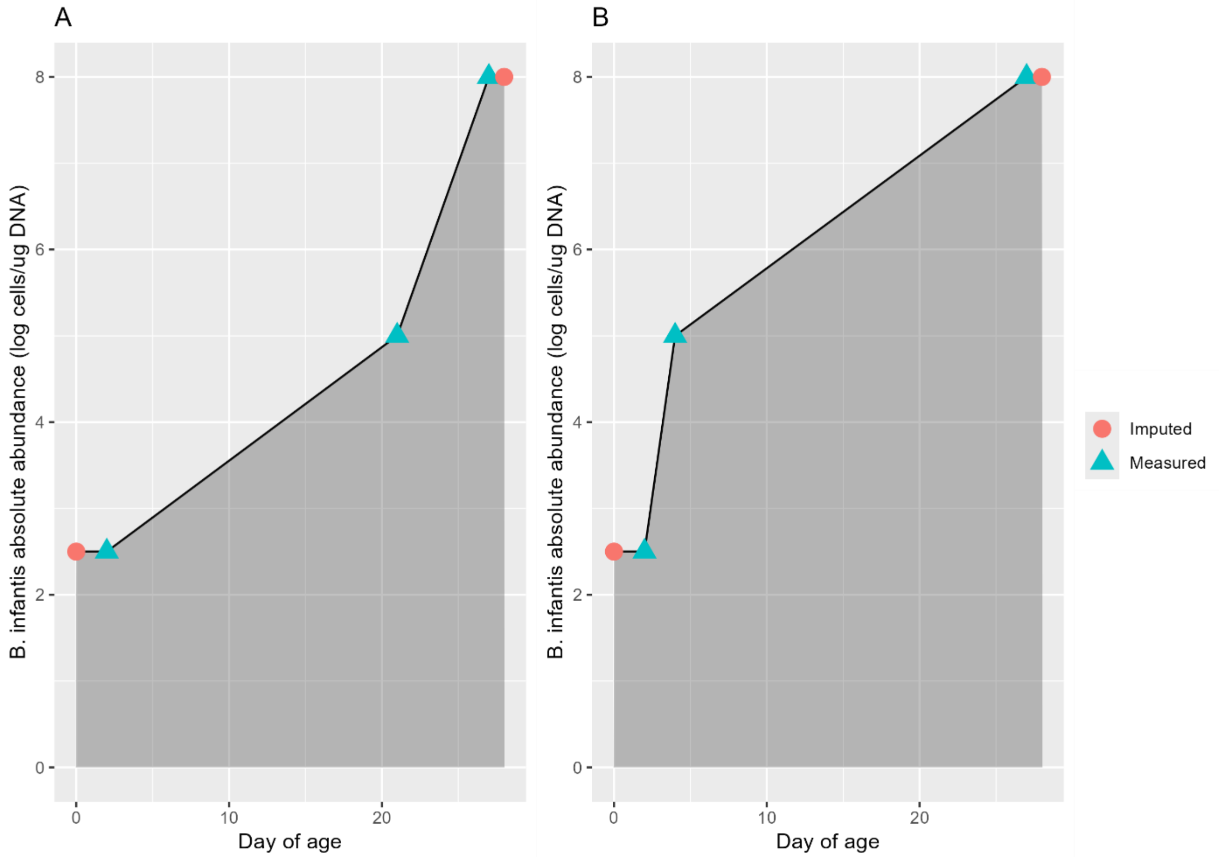

**Figure S1.** Calculation of *B. infantis* abundance area under the curve (AUC) for two example infants. Infants had stool samples with available *B. infantis* qPCR data at 4, 21, and 25 days of age (infant A), and at 4, 8, and 25 days of age (infant B). *B. infantis* qPCR data were unavailable at 0 and 28 days of age and were therefore imputed on both days for both infants.

In this example, the means and medians of infants A and B are the same but their AUCs differ slightly (**Table S1**). The mean and median are expressed in the same units as each single value, but the AUC has a less intuitive unit, which incorporates time-age axis (**Table S1**).

**Table S1.** Comparison of AUC, mean, and median between two example infants.

| Summary measure | Units | Infant A | Infant B |
|---|---|---|---|
| AUC | 100 log cells/µg DNA*days | 1.2 | 1.6 |
| Mean | log cells/µg DNA | 5.2 | 5.2 |
| Median | log cells/µg DNA | 5.0 | 5.0 |

Abbreviations: AUC, area under the curve.

In an analysis in which the summary measures were generated for all infants (n=966), all three measures (AUC, mean, and median) were strongly correlated with one another (**Figure S2A-C**). However, there was a relatively high proportion of median values corresponding to the imputed abundance value for samples with undetectable *B. infantis* (2.5 log cells/µg DNA) as well as at the highest measured abundances compared to AUC and mean values (**Figure S2, panels D-F**). The vertical columns of points on the scatterplots (panels A and C) show that the AUC and mean vary among infants with the same median value of 2.5 log cells/µg DNA (corresponding to the imputed abundance in one or more samples).

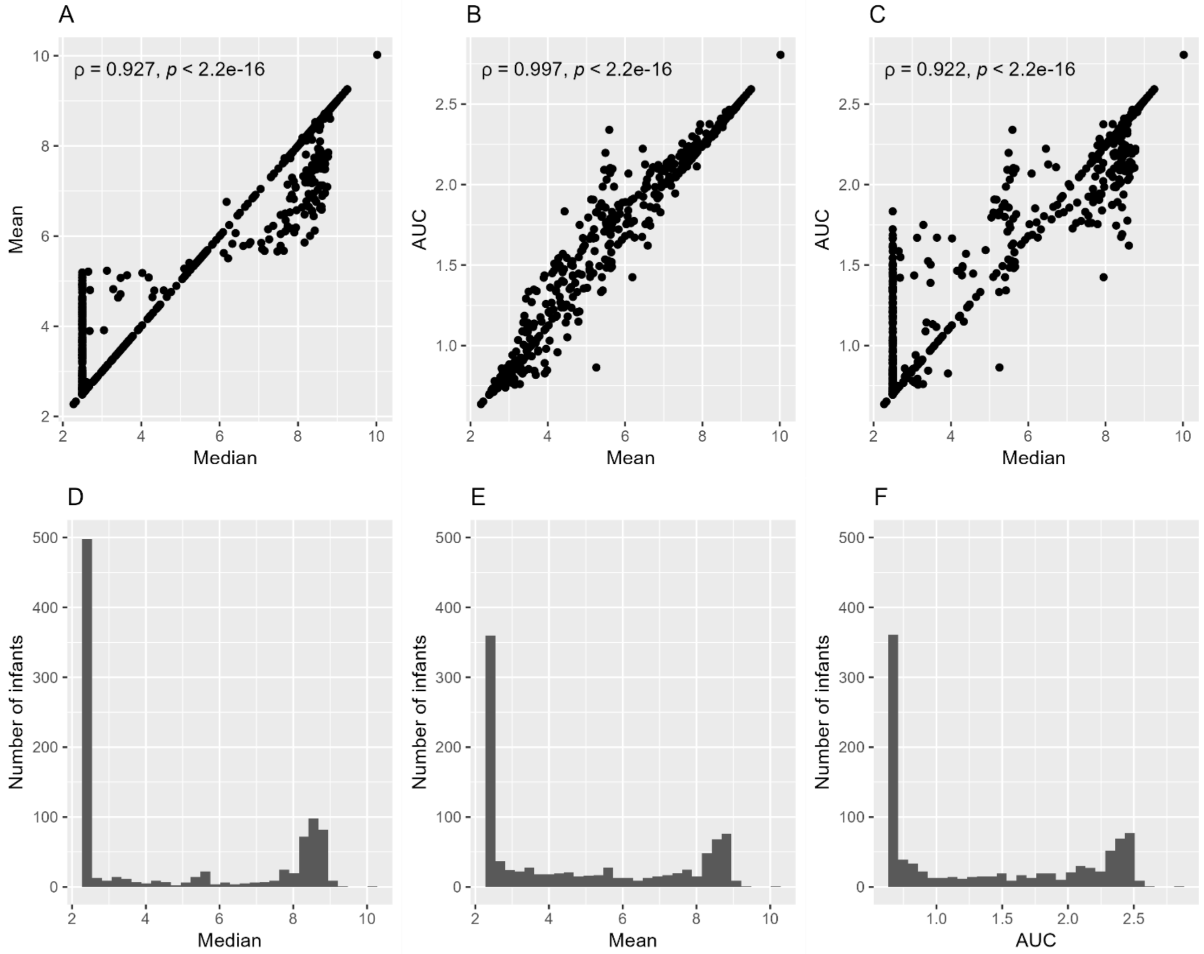

**Figure S2.** Comparison of mean versus median (A), AUC versus mean (B), and AUC versus median (C) of neonatal *B. infantis* absolute abundance. r, Spearman correlation coefficient. Distributions of the median (D), mean (E), and AUC (F) of neonatal *B. infantis* absolute abundance.
N samples=2951, N infants=966. Abbreviations: AUC, area under the curve.

To assess the robustness of each summary measure of *B. infantis* absolute abundance given a range of sample numbers per infant in the 0-28 day period, we progressively reduced the number of absolute abundance measurements used to generate the summary measure, among infants with 7 absolute abundance measurements. After successively omitting one randomly selected sample for each infant, abundance summary measures were generated based on the remaining samples. For each iteration, Spearman correlation coefficients were calculated across infants by comparing observed values in the reduced sample size to the corresponding summary measures based on all 7 samples (which served as the reference scenario). This process was repeated for 1000 iterations of random sampling. Mean Spearman correlations declined with successive removal of samples for all three measures (mean, median, and AUC), but were generally higher for AUC and mean, suggesting the median was less robust in the context of decreasing sample size (**Figure S3**).

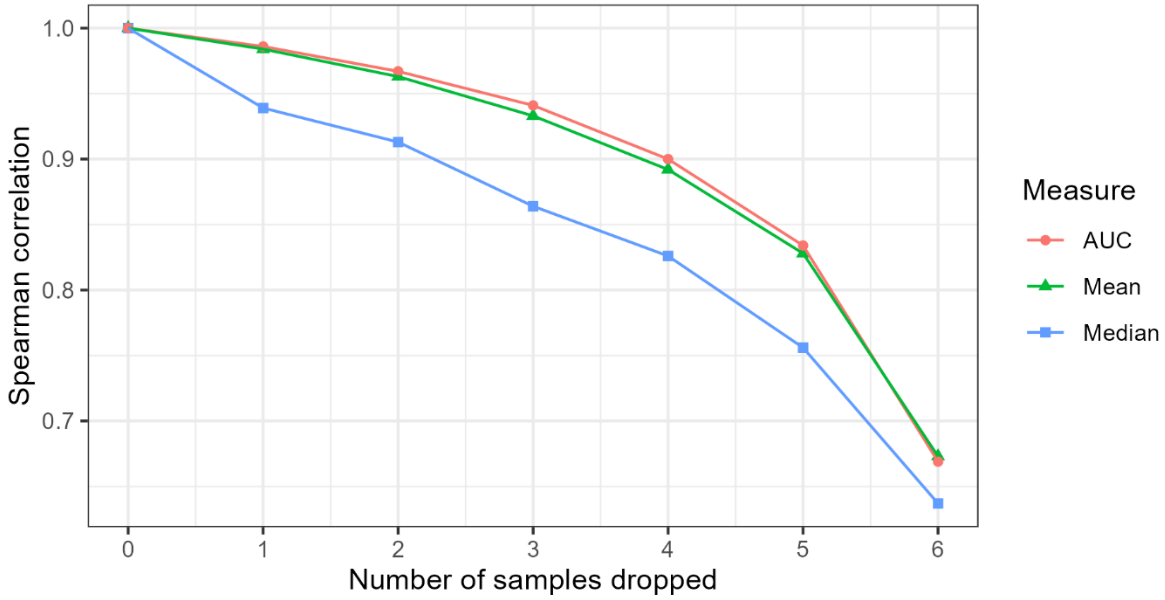

**Figure S3.** Mean Spearman correlations of infant-level *B. infantis* absolute abundance summary measures upon successive exclusion of up to 6 samples for each infant, based on 1000 iterations, among infants with 7 stool samples within 28 days after birth (N samples =1029, N infants =147). Abbreviations: AUC, area under the curve.

In the data analysis plan in the original protocol for this study, we specified the infant-level AUC of *B. infantis* abundance as the primary exposure variable. Our primary consideration was that the AUC incorporated information about the timing of sample collection and thereby may contribute to greater between-infant variability; however, based on the comparisons of the measures (per above), we selected mean *B. infantis* abundance to be the primary measure of *B. infantis* abundance as it performed empirically as well as AUC and has units that are easier to interpret (**Table S1**). Analyses were also conducted using AUCs, and the inferences were the same as those based on the means (primary results shown in **Table S10**).

### Supplementary Results

**Table S2.** Baseline characteristics of infants included in the primary analyses, stratified by infant sex.

| Characteristic | Median (25th, 75th centiles) or count (proportion) | |
|---|---|---|
| | Female (N=421) | Male (N=409) |
| Enrolment site | | |
| MFSTC | 242 (57) | 235 (57) |
| MCHTI | 179 (43) | 174 (43) |
| Baseline LAZ | -0.87 (-1.6, -0.23) | -0.78 (-1.4, -0.1) |
| Baseline WAZ[a] | -1.1 (-1.7, -0.48) | -0.96 (-1.6, -0.36) |
| Baseline WLZ[a] | -0.65 (-1.5, 0.12) | -0.58 (-1.3, 0.16) |
| Birth weight[a], kg | 2.80 (2.54, 3.03) | 2.94 (2.69, 3.20) |
| Mode of delivery | | |
| Vaginal | 208 (49) | 192 (47) |
| C-section | 213 (51) | 217 (53) |
| Stool collection schedule | | |
| A/B | 159 (38) | 161 (39) |
| C | 262 (62) | 248 (61) |
| Breastfeeding status by 28 days of age | | |
| Exclusively breastfed | 296 (70) | 265 (65) |
| Predominantly breastfed | 73 (17) | 84 (21) |
| Partially breastfed or not breastfed | 52 (12) | 60 (15) |
| Gestational age at birth | | |
| Term (≥260 days) | 394 (94) | 377 (92) |
| Preterm (<260 days) | 27 (6.4) | 32 (7.8) |
| Antibiotic use by 28 days of age | | |
| Yes | 49 (12) | 45 (11) |
| No | 372 (88) | 364 (89) |
| Maternal BMI, kg/m$^2$ | 26 (23, 29) | 26 (23, 29) |
| Maternal education | | |
| Little to no schooling | 110 (26) | 115 (28) |
| Secondary incomplete | 168 (40) | 137 (33) |
| Secondary complete or higher | 143 (34) | 157 (38) |
| Maternal age (years) | 24 (20, 28) | 24 (20, 28) |
| Number of infants within the household under five years of age | 0 (0, 1) | 0 (0, 1) |
| Household asset index quintile | | |
| 1 (lowest wealth) | 86 (20) | 67 (16) |
| 2 | 82 (19) | 86 (21) |
| 3 | 86 (20) | 80 (20) |
| 4 | 90 (21) | 89 (22) |
| 5 (highest wealth) | 77 (18) | 87 (21) |
| Number of infants with stool *B. infantis* absolute abundance data by 28 days of age | | |
| Infants with 1 sample | 172 (41) | 174 (43) |
| Infants with 2-4 samples | 102 (24) | 87 (21) |
| Infants with 5-7 samples | 147 (35) | 148 (36) |

Abbreviations: BMI, body mass index; LAZ, length-for-age z-score; MCHTI, Maternal and Child Health Training Institute; MFSTC, Mohammadpur Fertility Services and Training Centre; qPCR, quantitative polymerase chain reaction; WAZ, weight-for-age z-score; WLZ, weight-for-length z-score

[a] n female < 421 or n male < 409: baseline LAZ, n male=408; baseline WLZ, female=388 and n male=392. Baseline WAZ (n female=421, n male=409) and WLZ were calculated from weights measured at enrolment by study staff whereas birthweight (n female=421, n male=409) were extracted from the hospital records.

**Table S3.** Infant growth metrics in the overall SEPSiS observational cohort and this sub-study.

| Index | Age | SEPSiS observational cohort | | | *B. infantis* and growth sub-study[a] | | |
|---|---|---|---|---|---|---|---|
| | | **N infants** | **Mean** | **SD** | **N infants** | **Mean** | **SD** |
| LAZ | Baseline | 1841 | -0.80 | 1.0 | 829 | -0.81 | 0.99 |
| | 2 months | 1342 | -0.85 | 1.0 | 652 | -0.81 | 0.99 |
| | 3 months | 1353 | -0.72 | 1.0 | 677 | -0.72 | 1.0 |
| | 6 months | 1061 | -0.76 | 1.0 | 560 | -0.79 | 0.96 |
| WAZ | Baseline | 1840 | -0.99 | 0.93 | 830 | -1.0 | 0.95 |
| | 2 months | 1346 | -0.90 | 0.94 | 653 | -0.87 | 0.93 |
| | 3 months | 1359 | -0.77 | 1.0 | 680 | -0.77 | 0.95 |
| | 6 months | 1071 | -0.61 | 1.0 | 565 | -0.65 | 1.0 |
| WLZ | Baseline | 1755 | -0.59 | 1.2 | 780 | -0.61 | 1.2 |
| | 2 months | 1339 | -0.16 | 1.1 | 609 | -0.18 | 1.1 |
| | 3 months | 1350 | -0.23 | 1.0 | 634 | -0.25 | 1.1 |
| | 6 months | 1061 | -0.10 | 1.1 | 525 | -0.12 | 1.1 |

Abbreviations: LAZ, length-for-age z-score; WAZ, weight-for-age z-score; WLZ, weight-for-length z-score.

[a] Infants included in the primary analyses of the associations of neonatal *B. infantis* abundance and anthropometric outcomes.

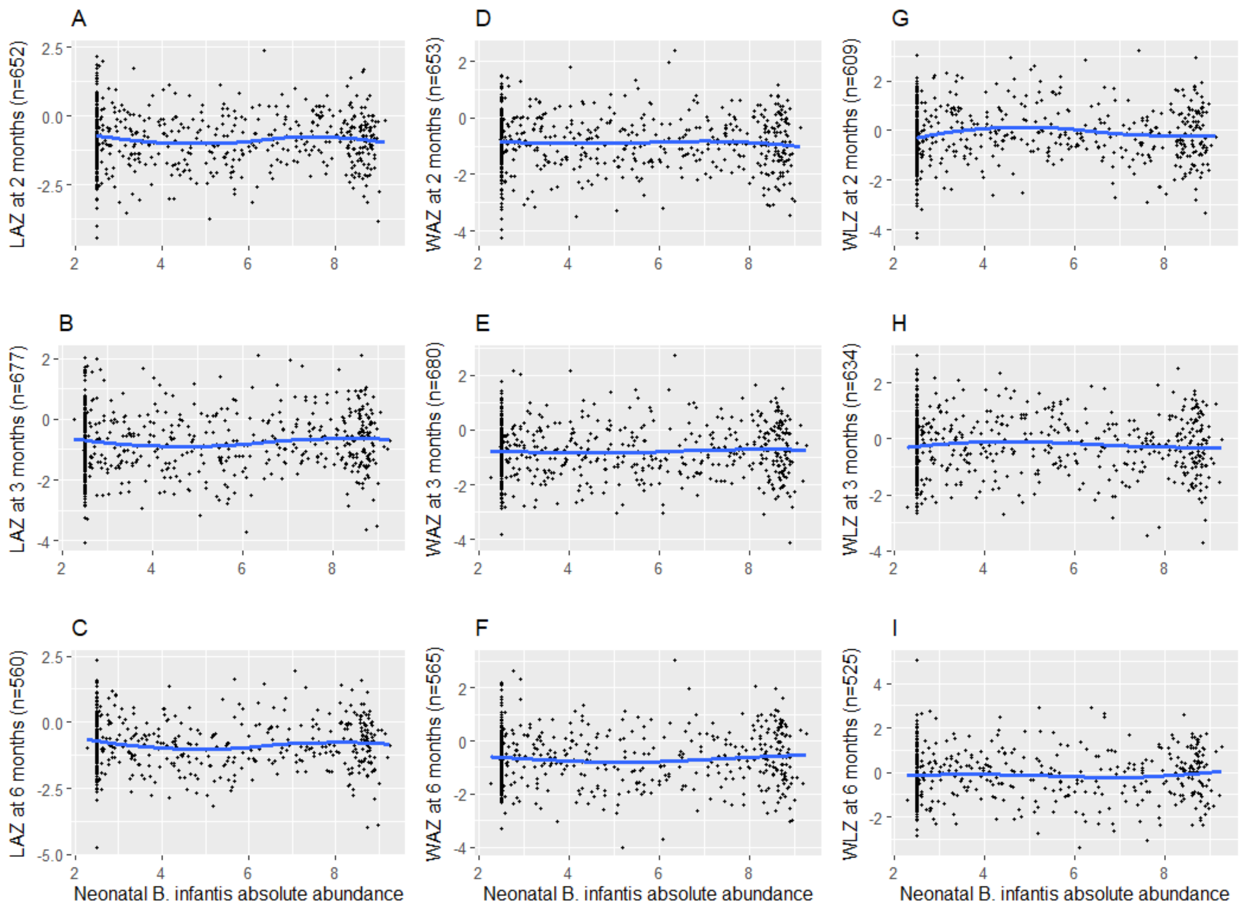

**Figure S4.** Relationships between infant anthropometric indices (LAZ: panels A-C, WAZ: panels D-F, and WLZ: panels G-I) at 2, 3, and 6 months of age and neonatal *B. infantis* absolute abundance up to 28 days of age, represented as loess curve (blue solid line).

Abbreviations: LAZ, length-for-age z-score; WAZ, weight-for-age z-score; WLZ, weight-for-length z-score.

**Table S4.** Associations between baseline anthropometric indices and neonatal *B. infantis* absolute abundance.

| Measure | Unadjusted model | | | Adjusted model[a] | | |
|---|---|---|---|---|---|---|
| | n | Difference in log cells/µg DNA of *B. infantis* absolute abundance[b] per 1 unit increase in anthropometric measure | p-value | n | Difference in log cells/µg DNA of *B. infantis* absolute abundance[b] per 1 unit increase in anthropometric measure | p-value |
| LAZ | 829 | -0.026 (-0.20, 0.15) | 0.8 | 829 | -0.013 (-0.18, 0.15) | 0.9 |
| WAZ | 830 | 0.091 (-0.088, 0.27) | 0.3 | 830 | 0.067 (-0.11, 0.25) | 0.5 |
| WLZ | 780 | 0.078 (-0.069, 0.22) | 0.3 | 780 | 0.078 (-0.064, 0.22) | 0.3 |

Abbreviations: AUC, area under the curve; CI, confidence interval; LAZ, length-for-age z-score; WAZ, weight-for-age z-score; WLZ, weight-for-length z-score.

[a] Covariates in adjusted models: enrolment site, infant sex, mode of delivery, gestational age at birth, maternal BMI, maternal education, maternal age, number of infants within the household under 5 years of age, and household asset index quintile.

[b] Neonatal *B. infantis* absolute abundance was defined at the individual infant level as the mean of the infant's *B. infantis* qPCR abundance data from all analyzed stool samples collected from 0-28 days (as described in the main text).

**Table S5**. Associations of neonatal *B. infantis* absolute abundance and anthropometric indices up to 2, 3, and 6 months of age, stratified by infant sex.

| Measure and age | Male infants | | | | | | Female infants | | | | | |
|---|---|---|---|---|---|---|---|---|---|---|---|---|
| | Unadjusted model[a] | | | Adjusted model[b] | | | Unadjusted model[a] | | | Adjusted model[b] | | |
| | n | Difference in anthropometric z-score (95% CI) for each 1 log cell/μg DNA increase in neonatal *B. infantis* absolute abundance [c] | p-value | n | Difference in anthropometric z-score (95% CI) for each 1 log cell/μg DNA increase in neonatal *B. infantis* absolute abundance [c] | p-value | n | Difference in anthropometric z-score (95% CI) for each 1 log cell/μg DNA increase in neonatal *B. infantis* absolute abundance [c] | p-value | n | Difference in anthropometric z-score (95% CI) for each 1 log cell/μg DNA increase in neonatal *B. infantis* absolute abundance [c] | p-value |
| LAZ | | | | | | | | | | | | |
| 2 months | 318 | -0.017 (-0.060, 0.025) | 0.4 | 318 | -0.011 (-0.051, 0.029) | 0.6 | 334 | -0.0082 (-0.043, 0.027) | 0.6 | 334 | 0.0083 (-0.025, 0.042) | 0.6 |
| 3 months | 338 | 0.0041 (-0.036, 0.044) | 0.8 | 338 | 0.0054 (-0.034, 0.044) | 0.8 | 339 | 0.014 (-0.022, 0.051) | 0.4 | 339 | 0.039 (0.0022, 0.075) | 0.04[d] |
| 6 months | 272 | -0.015 (-0.059, 0.028) | 0.5 | 272 | -0.0093 (-0.053, 0.035) | 0.7 | 288 | -0.0065 (-0.047, 0.034) | 0.8 | 288 | 0.029 (-0.011, 0.070) | 0.2 |
| WAZ | | | | | | | | | | | | |
| 2 months | 320 | -0.021 (-0.061, 0.018) | 0.3 | 320 | -0.026 (-0.066, 0.014) | 0.2 | 333 | -0.0017 (-0.036, 0.033) | >0.9 | 333 | 0.011 (-0.022, 0.045) | 0.5 |
| 3 months | 337 | -0.010 (-0.051, 0.031) | 0.6 | 337 | -0.012 (-0.054, 0.031) | 0.6 | 343 | 0.017 (-0.016, 0.051) | 0.3 | 343 | 0.027 (-0.0081, 0.062) | 0.1 |
| 6 months | 275 | -0.017 (-0.068, 0.034) | 0.5 | 275 | -0.024 (-0.076, 0.029) | 0.4 | 290 | 0.014 (-0.026, 0.054) | 0.5 | 290 | 0.024 (-0.019, 0.067) | 0.3 |
| WLZ | | | | | | | | | | | | |
| 2 months | 304 | -0.0082 (-0.060, 0.044) | 0.8 | 304 | -0.027 (-0.084, 0.029) | 0.3 | 305 | 0.015 (-0.035, 0.064) | 0.6 | 305 | 0.0054 (-0.047, 0.058) | 0.8 |
| 3 months | 323 | -0.023 (-0.070, 0.025) | 0.3 | 323 | -0.030 (-0.080, 0.021) | 0.2 | 311 | 0.0039 (-0.042, 0.049) | 0.9 | 311 | -0.014 (-0.063, 0.034) | 0.6 |
| 6 months | 260 | -0.026 (-0.082, 0.031) | 0.4 | 260 | -0.039 (-0.098, 0.020) | 0.2 | 265 | 0.028 (-0.026, 0.081) | 0.3 | 265 | 0.0023 (-0.054, 0.059) | >0.9 |

Abbreviations: CI, confidence interval; LAZ, length-for-age z-score; WAZ, weight-for-age z-score; WLZ, weight-for-length z-score.

[a]Baseline (enrolment) measurement of the same parameter as the outcome was included as a covariate in all models.

[b]Additional covariates in adjusted models: enrolment site, infant sex, breastfeeding status at 28 days of age (if available) or at age most proximal to 28 days, mode of delivery, gestational age at birth, infant antibiotic exposure by 28 days of age, maternal BMI, maternal education, maternal age, number of infants within the household under 5 years of age, and household asset index quintile.

[c] Neonatal *B. infantis* absolute abundance was defined at the individual infant level as the mean of the infant's *B. infantis* qPCR abundance data from all analyzed stool samples collected from 0-28 days (as described in the main text).

[d] No longer statistically significant after Holm's procedure.

**Table S6.** Sensitivity analysis: associations of dichotomous neonatal *B. infantis* absolute abundance (detectable vs. not detectable) and anthropometric indices at 2, 3, and 6 months of age.

| Measure and age | Unadjusted model[a] | | | Adjusted model[b] | | |
|---|---|---|---|---|---|---|
| | n | Difference in anthropometric z-score (95% CI) between infants with detectable *B. infantis* compared to infants with no detectable *B. infantis* | p-value | n | Difference in anthropometric z-score (95% CI) between infants with detectable *B. infantis* compared to infants with no detectable *B. infantis* | p-value |
| LAZ | | | | | | |
| 2 months | 652 | -0.12 (-0.26, 0.020) | 0.09 | 652 | -0.062 (-0.19, 0.061) | 0.3 |
| 3 months | 677 | -0.023 (-0.16, 0.12) | 0.7 | 677 | 0.035 (-0.092, 0.16) | 0.6 |
| 6 months | 560 | -0.13 (-0.29, 0.016) | 0.08 | 560 | -0.050 (-0.19, 0.094) | 0.5 |
| WAZ | | | | | | |
| 2 months | 653 | -0.023 (-0.16, 0.11) | 0.7 | 653 | 0.042 (-0.088, 0.17) | 0.5 |
| 3 months | 680 | -0.011 (-0.15, 0.12) | 0.9 | 680 | 0.059 (-0.077, 0.19) | 0.4 |
| 6 months | 565 | -0.028 (-0.19, 0.14) | 0.7 | 565 | 0.028 (-0.14, 0.19) | 0.7 |
| WLZ | | | | | | |
| 2 months | 609 | 0.18 (0.0000 0.37) | 0.05[c] | 609 | 0.15 (-0.038, 0.33) | 0.1 |
| 3 months | 634 | 0.015 (-0.15, 0.18) | 0.9 | 634 | 0.018 (-0.15, 0.19) | 0.8 |
| 6 months | 525 | 0.046 (-0.15, 0.24) | 0.6 | 525 | 0.025 (-0.18, 0.23) | 0.8 |

Abbreviations: CI, confidence interval; LAZ, length-for-age z-score; WAZ, weight-for-age z-score; WLZ, weight-for-length z-score.

[a] Baseline (enrolment) measurement of the same parameter as the outcome was included as a covariate in all models.

[b] Additional covariates in adjusted models: enrolment site, infant sex, breastfeeding status at 28 days of age (if available) or at age most proximal to 28 days, mode of delivery, gestational age at birth, infant antibiotic exposure by 28 days of age, maternal BMI, maternal education, maternal age, number of infants within the household under 5 years of age, and household asset index quintile.

[c] Value rounded up from below 0.05. No longer statistically significant after Holm's procedure.

**Table S7.** Sensitivity analysis: associations of mean *B. infantis* absolute abundance up to 14 days of age and anthropometric indices at 2, 3, and 6 months of age.

| Measure | Age (months) | n[a] | Difference in anthropometric z-score (95% CI) for each 1 log cell/µg DNA increase in mean *B. infantis* absolute abundance[b,c] | p-value |
|---|---|---|---|---|
| LAZ | 2 | 608 | 0.0054 (-0.021, 0.032) | 0.7 |
| | 3 | 625 | 0.018 (-0.010, 0.046) | 0.2 |
| | 6 | 527 | 0.0063 (-0.025, 0.038) | 0.7 |
| WAZ | 2 | 608 | -0.0082 (-0.035, 0.019) | 0.6 |
| | 3 | 627 | 0.0062 (-0.023, 0.035) | 0.7 |
| | 6 | 532 | 0.0076 (-0.028, 0.044) | 0.7 |
| WLZ | 2 | 567 | -0.020 (-0.060, 0.021) | 0.3 |
| | 3 | 583 | -0.020 (-0.058, 0.018) | 0.3 |
| | 6 | 493 | -0.0060 (-0.050, 0.038) | 0.8 |

Abbreviations: CI, confidence interval; LAZ, length-for-age z-score; WAZ, weight-for-age z-score; WLZ, weight-for-length z-score.

[a] N is lower than in the primary analyses because fewer infants had stool samples collected at 0-14 than 0-28 days of age.

[b] Covariates in adjusted model include baseline measurement of the same parameter as the outcome at enrolment, enrolment site, infant sex, breastfeeding status as near to day 14 as possible, mode of delivery, gestational age at birth, infant antibiotics use by 14 days of age, maternal BMI, maternal education, maternal age, the number of infants within the household 5 five years of age, and household asset index quintile.

[c] Neonatal *B. infantis* absolute abundance was defined at the individual infant level as the mean of the infant's *B. infantis* qPCR abundance data from all analyzed stool samples collected from 0-14 days.

**Table S8.** Sensitivity analysis: associations of mean *B. infantis* absolute abundance up to 60 days of age and anthropometric indices at 2, 3, and 6 months of age.

| Measure | Age (months) | n | Difference in anthropometric z-score (95% CI) for each 1 log cell/µg DNA increase in mean *B. infantis* absolute abundance[a,b] | p-value |
|---|---|---|---|---|
| LAZ | 2 | 764 | -0.020 (-0.044, 0.0037) | 0.1 |
| | 3 | 763 | 0.0054 (-0.020, 0.031) | 0.7 |
| | 6 | 631 | -0.0052 (-0.033, 0.023) | 0.7 |
| WAZ | 2 | 766 | -0.018 (-0.041, 0.0062) | 0.1 |
| | 3 | 767 | -0.0016 (-0.028, 0.025) | >0.9 |
| | 6 | 637 | -0.0030 (-0.035, 0.029) | 0.9 |
| WLZ | 2 | 715 | -0.0016 (-0.036, 0.033) | >0.9 |
| | 3 | 717 | -0.011 (-0.045, 0.022) | 0.5 |
| | 6 | 592 | -0.00056 (-0.039, 0.038) | >0.9 |

Abbreviations: CI, confidence interval; LAZ, length-for-age z-score; WAZ, weight-for-age z-score; WLZ, weight-for-length z-score.

[a] Covariates in adjusted model include baseline measurement of the same parameter as the outcome at enrolment, enrolment site, infant sex, breastfeeding status as near to 28 days of age as possible, mode of delivery, gestational age at birth, infant antibiotics use by 28 days of age, maternal BMI, maternal education, maternal age, the number of infants within the household under 5 years of age, and household asset index quintile. Inferences remained unchanged if breastfeeding status and antibiotics use were extended to 60 days of age.

[b] Neonatal *B. infantis* absolute abundance was defined at the individual infant level as the mean of the infant's *B. infantis* qPCR abundance data from all analyzed stool samples collected from 0-60 days.

**Table S9.** Sensitivity analysis: cross-sectional associations between *B. infantis* absolute abundance measured in a single stool sample collected closest in time to the 2-month anthropometric measurement.

| Measure | Unadjusted model[a] | | | Adjusted model[b] | | |
|---|---|---|---|---|---|---|
| | n | Difference in anthropometric z-score per increase in 1 log cells/µg DNA of *B. infantis* (95% CI) | p-value | n | Difference in anthropometric z-score per increase in 1 log cells/µg DNA of *B. infantis* (95% CI) | p-value |
| LAZ | 393 | 0.0059 (-0.029, 0.041) | 0.7 | 393 | 0.0043 (-0.027, 0.036) | 0.8 |
| WAZ | 396 | 0.013 (-0.020, 0.046) | 0.4 | 396 | 0.0052 (-0.026, 0.036) | 0.7 |
| WLZ | 369 | 0.0063 (-0.037, 0.050) | 0.8 | 369 | -0.0073 (-0.052, 0.037) | 0.7 |

Abbreviations: CI, confidence interval; LAZ, length-for-age z-score; WAZ, weight-for-age z-score; WLZ, weight-for-length z-score.

[a] Baseline (enrolment) measurement of the same parameter as the outcome was included as a covariate in all models.

[b] Covariates in adjusted model include baseline measurement of the same parameter as the outcome at enrolment, enrolment site, infant sex, breastfeeding status at 28 days of age (if available) or at age most proximal to 28 days, mode of delivery, gestational age at birth, infant antibiotic exposure by 28 days of age, maternal BMI, maternal education, maternal age, the number of infants within the household under 5 years of age, and household asset index quintile.

**Table S10.** Sensitivity analysis: Associations between *B. infantis* absolute abundance area under the curve (0-28 days of age) and infant anthropometric indices at 2, 3, and 6 months of age.

| Measure and age | Unadjusted model[a] | | | Adjusted model[b] | | |
|---|---|---|---|---|---|---|
| | n | Difference in anthropometric z-score (95% CI) for each 1 unit of area under the curve of *B. infantis* | p-value | n | Difference in anthropometric z-score (95% CI) for each 1 unit of area under the curve of *B. infantis* | p-value |
| LAZ | | | | | | |
| 2 months | 652 | -0.051 (-0.15, 0.044) | 0.3 | 652 | -0.0090 (-0.098, 0.080) | 0.8 |
| 3 months | 677 | 0.036 (-0.058, 0.13) | 0.5 | 677 | 0.079 (-0.013, 0.17) | 0.09 |
| 6 months | 560 | -0.055 (-0.16, 0.049) | 0.3 | 560 | 0.016 (-0.087, 0.12) | 0.8 |
| WAZ | | | | | | |
| 2 months | 653 | -0.041 (-0.13, 0.049) | 0.4 | 653 | -0.025 (-0.11, 0.064) | 0.6 |
| 3 months | 680 | 0.016 (-0.076, 0.11) | 0.7 | 680 | 0.034 (-0.061, 0.13) | 0.5 |
| 6 months | 565 | -0.017 (-0.13, 0.096) | 0.8 | 565 | -0.0073 (-0.12, 0.11) | >0.9 |
| WLZ | | | | | | |
| 2 months | 609 | 0.034 (-0.092, 0.16) | 0.6 | 609 | -0.021 (-0.15, 0.11) | 0.8 |
| 3 months | 634 | -0.030 (-0.14, 0.084) | 0.6 | 634 | -0.065 (-0.19, 0.056) | 0.3 |
| 6 months | 525 | 0.0048 (-0.13, 0.14) | >0.9 | 525 | -0.056 (-0.20, 0.086) | 0.4 |

Abbreviations: CI, confidence interval; LAZ, length-for-age z-score; WAZ, weight-for-age z-score; WLZ, weight-for-length z-score.

[a] Baseline (enrolment) measurement of the same parameter as the outcome was included as a covariate in all models.

[b] Additional covariates in adjusted models: enrolment site, infant sex, breastfeeding status at 28 days of age (if available) or at age most proximal to 28 days, mode of delivery, gestational age at birth, infant antibiotic exposure by 28 days of age, maternal BMI, maternal education, maternal age, number of infants within the household under 5 years of age, and household asset index quintile.

**Table S11.** Sensitivity analysis: Associations of neonatal *B. infantis* absolute abundance and infant anthropometric indices at 2, 3, and 6 months of age without adjustment for baseline anthropometric measures.

| Measure and age | Unadjusted model | | | Adjusted model[a] | | |
|---|---|---|---|---|---|---|
| | n | Difference in anthropometric z-score (95%CI) per 1 unit increase in log cells/µg DNA of neonatal *B. infantis* abundance[b] | p-value | n | Difference in anthropometric z-score (95%CI) per 1 unit increase in log cells/µg DNA of neonatal *B. infantis* abundance[b] | p-value |
| LAZ | | | | | | |
| 2 months | 652 | -0.015 (-0.046, 0.016) | 0.4 | 652 | -0.00075 (-0.033, 0.032) | >0.9 |
| 3 months | 677 | 0.013 (-0.018, 0.044) | 0.4 | 677 | 0.028 (-0.0046, 0.060) | 0.09 |
| 6 months | 560 | -0.0079 (-0.040, 0.024) | 0.6 | 560 | 0.011 (-0.022, 0.045) | 0.5 |
| WAZ | | | | | | |
| 2 months | 653 | -0.0063 (-0.036, 0.023) | 0.7 | 653 | -0.0040 (-0.034, 0.026) | 0.8 |
| 3 months | 680 | 0.014 (-0.016, 0.043) | 0.4 | 680 | 0.017 (-0.013, 0.048) | 0.3 |
| 6 months | 565 | 0.0058 (-0.028, 0.040) | 0.7 | 565 | 0.0080 (-0.028, 0.044) | 0.7 |
| WLZ | | | | | | |
| 2 months | 609 | 0.0083 (-0.028, 0.045) | 0.7 | 609 | -0.0085 (-0.047, 0.030) | 0.7 |
| 3 months | 634 | -0.0073 (-0.041, 0.026) | 0.7 | 634 | -0.018 (-0.053, 0.018) | 0.3 |
| 6 months | 525 | 0.0024 (-0.037, 0.042) | >0.9 | 525 | -0.014 (-0.055, 0.028) | 0.5 |

Abbreviations: CI, confidence interval; LAZ, length-for-age z-score; WAZ, weight-for-age z-score; WLZ, weight-for-length z-score.

[a] Covariates in adjusted models: enrolment site, infant sex, breastfeeding status at 28 days of age (if available) or at age most proximal to 28 days, mode of delivery, gestational age at birth, infant antibiotic exposure by 28 days of age, maternal BMI, maternal education, maternal age, number of infants within the household under 5 years of age, and household asset index quintile.

[b] Neonatal *B. infantis* absolute abundance was defined at the individual infant level as the mean of the infant's *B. infantis* qPCR abundance data from all analyzed stool samples collected from 0-28 days (as described in the main text).

**Table S12.** Associations between neonatal *B. infantis* absolute abundance and infant anthropometric indices up to 2, 3, and 6 months of age in analyses restricted to infants in schedule A/B.

| Measure | Age (months) | n | Difference in anthropometric z-score per 1 unit increase in log cells of *B. infantis*/µg DNA per day (95%CI)[a,b] | p-value |
|---|---|---|---|---|
| LAZ | 2 | 263 | 0.073 (0.020, 0.13) | 0.007[c] |
| | 3 | 269 | 0.087 (0.032, 0.14) | 0.002[d] |
| | 6 | 231 | 0.033 (-0.031, 0.096) | 0.3 |
| WAZ | 2 | 265 | -0.0062 (-0.058, 0.045) | 0.8 |
| | 3 | 269 | 0.0098 (-0.051, 0.071) | 0.8 |
| | 6 | 234 | 0.0036 (-0.077, 0.084) | >0.9 |
| WLZ | 2 | 249 | -0.12 (-0.20, -0.042) | 0.003[d] |
| | 3 | 254 | -0.11 (-0.19, -0.029) | 0.008[c] |
| | 6 | 220 | -0.056 (-0.15, 0.038) | 0.2 |

Abbreviations: CI, confidence interval; LAZ, length-for-age z-score; WAZ, weight-for-age z-score; WLZ, weight-for-length z-score.

[a] Covariates in adjusted models: baseline (enrolment) measurement of the same parameter as the outcome, enrolment site, infant sex, breastfeeding status at 28 days of age (if available) or at age most proximal to 28 days, mode of delivery, gestational age at birth, infant antibiotic exposure by 28 days of age, maternal BMI, maternal education, maternal age, number of infants within the household under 5 years of age, and household asset index quintile.

[b] Neonatal *B. infantis* absolute abundance was defined at the individual infant level as the mean of the infant's *B. infantis* qPCR abundance data from all analyzed stool samples collected from 0-28 days (as described in the main text).

[c] No longer statistically significant after Holm's procedure.

[d] Remained statistically significant after Holm's procedure (accounting for all analyses in this table).

**Table S13.** Associations of *B. infantis* absolute abundance up to 28 days of age (based on 2 randomly selected stool samples) and LAZ at 2, 3, and 6 months of age among infants in schedule A/B.

| Age (months) | n infants | Analysis based on 2 randomly selected abundance measures [a] | | Analysis based on all abundance measures, 0-28 days [b] | |
|---|---|---|---|---|---|
| | | Difference in LAZ per 1 unit increase in log cells of *B. infantis*/µg DNA per day (95%CI), mean (95% CI)[c] | p-value, mean (SD) | Difference in LAZ per 1 unit increase in log cells of *B. infantis*/µg DNA per day (95%CI), mean (95% CI)[c] | p-value |
| 2 | 246 | 0.054 (0.022, 0.085) | 0.1 (0.0034, 0.43) | 0.083 (0.027, 0.14) | 0.004 |
| 3 | 247 | 0.074 (0.042, 0.10) | 0.03 (0.00037, 0.16) | 0.091 (0.035, 0.15) | 0.002 |
| 6 | 208 | 0.043 (0.0060, 0.080) | 0.3 (0.022, 0.83) | 0.043 (-0.024, 0.11) | 0.2 |

Abbreviations: CI, confidence interval; LAZ, length-for-age z-score; WAZ, weight-for-age z-score; WLZ, weight-for-length z-score.

[a] One qPCR measurement was randomly selected from 0-4 days of age and the second sample was from 5-28 days of age. The mean of the two values was used in adjusted models. The mean (95% CI) of the coefficients and p values from 1000 iterations are shown.

[b] Abundance based on up to seven neonatal samples, but restricted to infants included in the analyses using only two randomly-selected samples.

[c] Covariates in adjusted models: baseline measurement (on day of enrolment) of the LAZ, enrolment site, infant sex, breastfeeding status at 28 days of age (if available) or at age most proximal to 28 days, mode of delivery, gestational age at birth, infant antibiotic exposure by 28 days of age, maternal BMI, maternal education, maternal age, number of infants within the household under 5 years of age, and household asset index quintile.

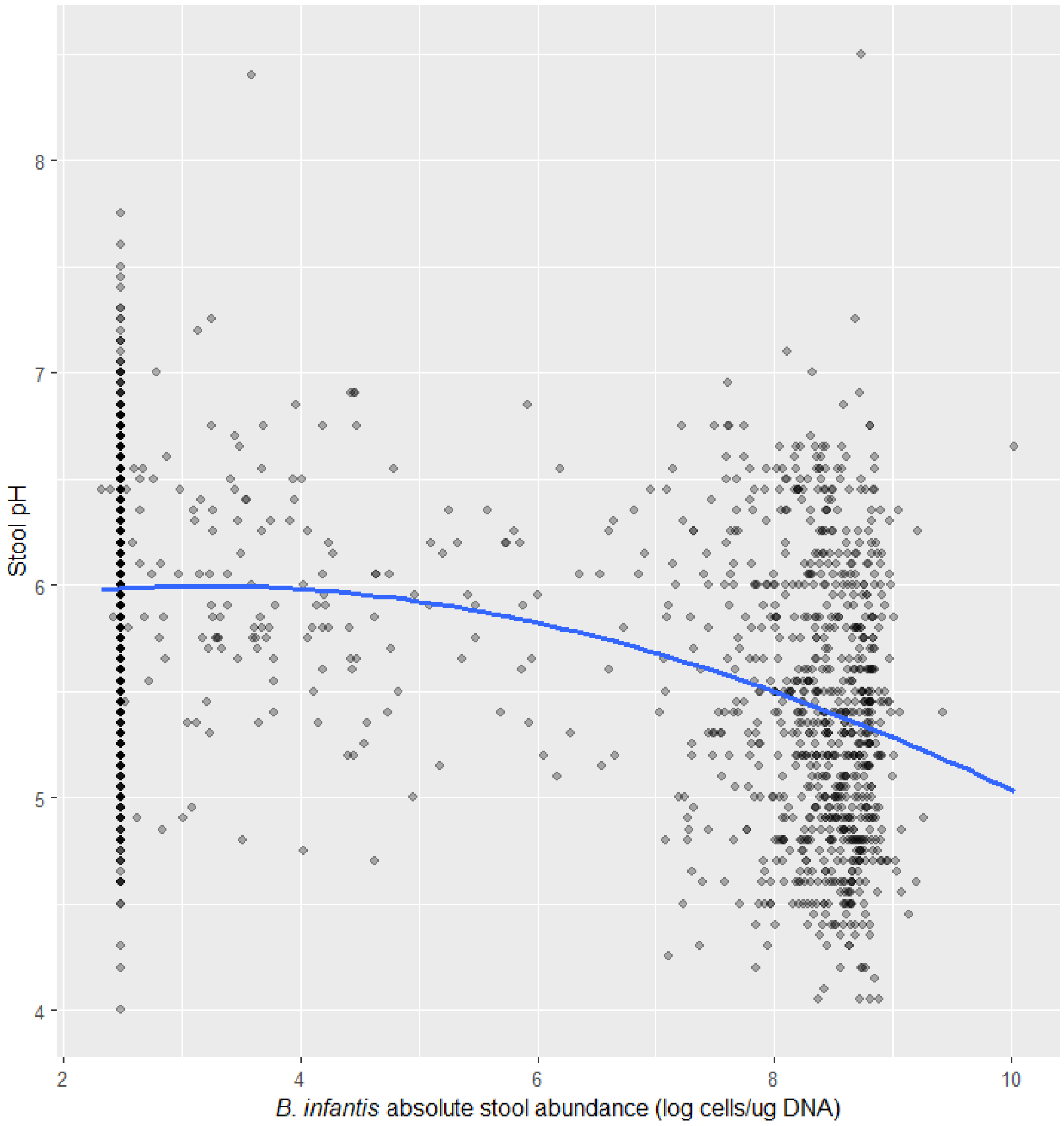

**Figure S5.** Stool pH by *B. infantis* stool absolute abundance in stool samples collected within the first 28 days after birth.

The blue fit line is a loess curve (N=845 infants; 2118 stool samples). Darker shading represents an overlap of points.

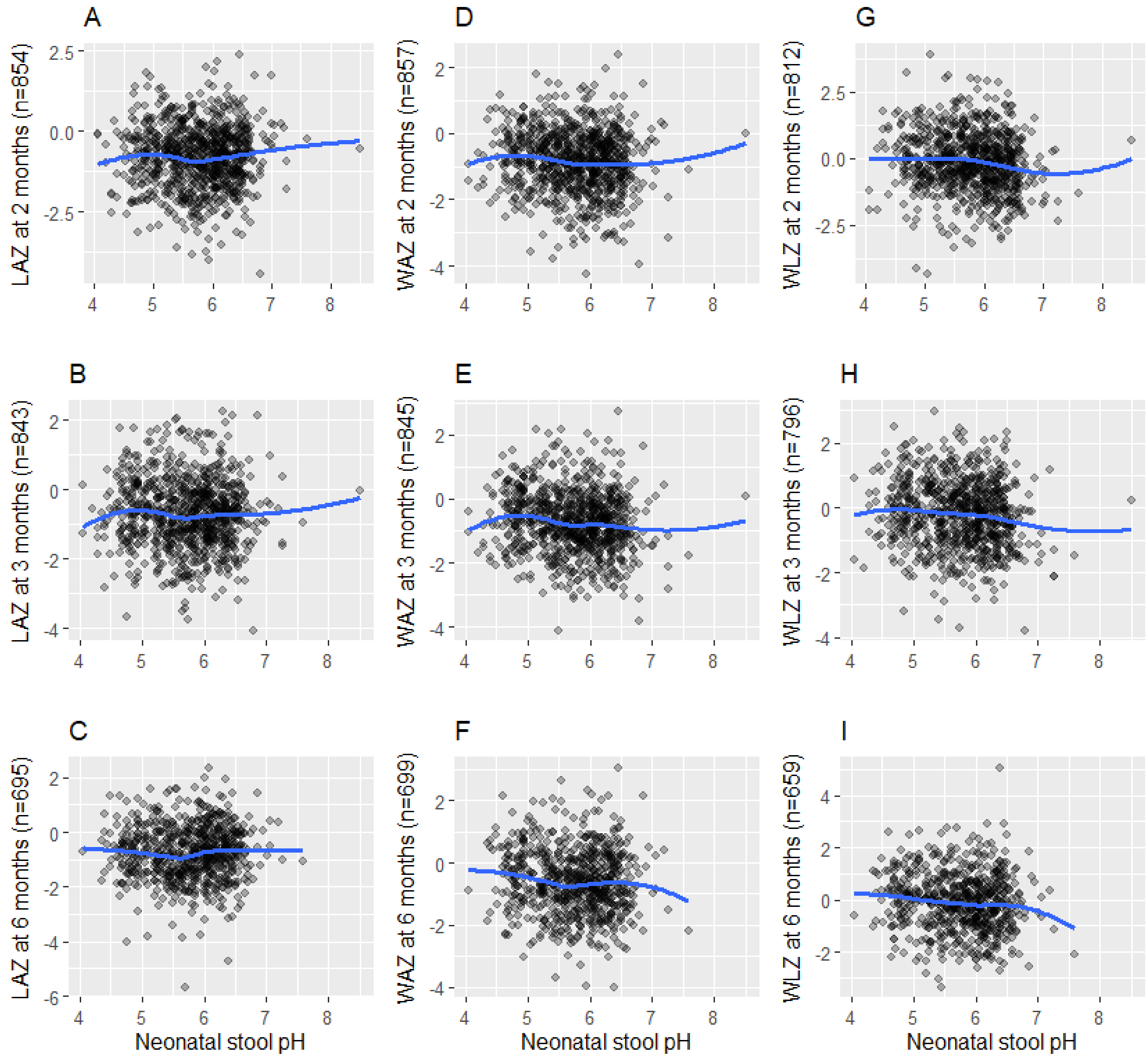

**Figure S6.** Associations between infant anthropometric indices at 2, 3, and 6 months of age and neonatal stool pH (LAZ: panels A-C; WAZ: panels D-F; WLZ: panels G-I).

Abbreviations: LAZ, length-for-age z-score; WAZ, weight-for-age z-score; WLZ, weight-for-length z-score.

**Table S14.** Sensitivity analyses: cross-sectional associations of stool pH and anthropometric indices at 2, 3, and 6 months of age.[a]

| Measure and age | n | Difference in anthropometric z-score per 1 unit decrease in pH (95%CI)[b] | p-value |
|---|---|---|---|
| LAZ | | | |
| 2 months | 395 | -0.074 (-0.20, 0.051) | 0.2 |
| 3 months | 248 | 0.098 (-0.073, 0.27) | 0.3 |
| 6 months | 188 | -0.048 (-0.23, 0.13) | 0.6 |
| WAZ | | | |
| 2 months | 396 | -0.0043 (-0.13, 0.12) | >0.9 |
| 3 months | 249 | 0.040 (-0.15, 0.23) | 0.7 |
| 6 months | 192 | -0.091 (-0.33, 0.15) | 0.5 |
| WLZ | | | |
| 2 months | 379 | 0.081 (-0.086, 0.25) | 0.3 |
| 3 months | 236 | -0.060 (-0.33, 0.21) | 0.7 |
| 6 months | 179 | 0.016 (-0.27, 0.30) | >0.9 |

Abbreviations: CI, confidence interval; LAZ, length-for-age z-score; WAZ, weight-for-age z-score; WLZ, weight-for-length z-score.

[a] These are cross-sectional analyses whereby the pH measure is the value from a single stool sample collected at the age that is closest in time to the day on which the infant's anthropometric measure was obtained, within the same period of 2 months (59-75 days), 3 months (76-110 days), and 6 months (160 – 200 days) of age.

[b] Adjusted model including the following covariates: baseline (enrolment) measurement of the same parameter as the outcome, enrolment site, infant sex, breastfeeding status as near to 28 days of age as possible, mode of delivery, gestational age at birth, antibiotics use by 28 days of age, maternal BMI, maternal education, maternal age, the number of infants within the household under 5 years of age, and household asset index quintile.
